# Changes in hierarchical brain dynamics of rumination following mindfulness-based cognitive therapy for depression

**DOI:** 10.64898/2026.06.21.26356048

**Authors:** Paulina Clara Dagnino, Anne Maj van der Velden, Henricus G. Ruhe, Willem Kuyken, Morten L. Kringelbach, Jakub Vohryzek, Gustavo Deco

## Abstract

Major depressive disorder (MDD) is a leading cause of disability worldwide with risk of onset and recurrence linked to depressive ruminative thought patterns. Mindfulness-based cognitive therapy (MBCT) is an evidence-based treatment for depression that targets the ability to recognise, decenter, and disengage from ruminative thought patterns. Elucidating how MBCT impacts hierarchical brain organisation may be key to understanding the processes by which MBCT can modulate ruminative tendencies. In a randomised controlled functional magnetic resonance imaging (fMRI) trial on individuals with MDD (N=80) before and after MBCT in addition to treatment as usual (TAU), we investigated changes in hierarchical brain organisation during resting-state and rumination. We built whole-brain models to obtain generative connectivity (GEC) matrices per patient and quantified brain hierarchy by measuring the global directedness and regional trophic levels in each GEC, in which greater directedness reflects more directional information flow and less recurrence. Global directedness in MBCT+TAU compared to TAU increased during rumination, with no changes during resting-state. Furthermore, increased regional breadth of hierarchy during rumination was related to improvements in clinical and behavioural outcomes following MBCT+TAU. Increased brain hierarchy during rumination following mindfulness training may be consistent with a shift away from self-reinforcing negative mental loops towards more differentiated and less coupled cognitive and bodily cycles, supporting MBCT’s ability to interrupt ruminative processes. Hierarchical brain dynamics may hold promise as a treatment-sensitive marker and a potential mechanism of therapeutic change in MBCT for depression.

**Trial registration:** ClinicalTrials.gov NCT03353493.

## Introduction

Major depressive disorder (MDD), one of the leading causes of global disability, is a complex mental disorder severely impairing functioning and quality of life (WHO, 2017). It is characterised by low mood, anhedonia, and the dominance and persistence of rumination, repetitive negative thought patterns that are linked to onset and recurrence of depression. An evidence-based and cost-effective treatment that targets this depressive vulnerability is mindfulness-based cognitive therapy (MBCT), which has shown similar efficacy and potential as an alternative to antidepressant medication (ADM) (Kuyken et al., 2016). MBCT consists of a group-delivered psychological intervention combining elements from cognitive behavioural therapy with mindfulness practice. It was initially designed to be delivered as a preventative treatment during remission to prevent relapse (Segal, 2002), but the evidence has since then expanded to show that MBCT can be also effective during acute and treatment resistant depression (Barnhofer et al., 2025; Goldberg et al., 2019; Pots et al., 2014; Thimm & Johnsen, 2020; Tickell et al., 2020; van Aalderen et al., 2015).

Initial episodes of depression are mainly associated to current stressful events, adversity in early life and genetic vulnerability (Beck, 2008). In recovered depressed patients, there is a vulnerability for small changes in mood to reactivate thinking styles associated with past episodes. These are driven by rumination, in which individuals dwell on negative problems and feelings in a belief that understanding their causes will help find solutions. This negative self-perpetuating cycle can then spiral down towards new depressive episodes. Furthermore, during depressive episodes, in treatment-resistant, or chronic depression, this cycle often becomes locked in a reinforcing loop maintaining depressive symptoms (Teasdale et al., 1995). In this view, depressive vulnerability could be reduced if patients learn to be more aware of thoughts and feelings and respond by disengaging from ruminative depressive processing (Teasdale et al., 2000). To this aim, MBCT targets vulnerability to depression by teaching the ability to recognise, decenter, and disengage from ruminative negative thoughts, and by fostering meta-awareness, a non-judgemental and compassionate attitude towards present moment experience.

To optimise MBCT it is very important to understand the psychological processes and brain mechanisms behind the therapy. With respect to brain markers, past investigations with functional magnetic resonance imaging (fMRI) have shown changes mainly in areas of the default mode network (DMN) and salience network (SAL) following mindfulness-based training (Tang, 2020; Tang et al., 2015; Vignaud et al., 2018; Young et al., 2018), but few studies have looked specifically at the neural mechanisms in MBCT for depression (van der Velden & Roepstorff, 2015; van der Velden, 2015). In our randomised controlled trial of MDD with MBCT in addition to treatment as usual (TAU) (MBCT+TAU), and TAU, fMRI scans were assessed before and after treatment during resting-state, mindfulness and an experimentally induced task of rumination (van der Velden, 2023). Our a priory static functional connectivity matrices showed decreased connectivity from the SAL to the lingual gyrus in MBCT+TAU compared to TAU during rumination and not resting-state nor mindfulness (van der Velden, 2023). This change mediated improvements in the ability to control and sustain bodily sensation attention. Moreover, a data-driven network analysis during rumination showed a reduction in a phase-locking pattern led by areas mainly from the SAL and somatomotor network (SOM) following MBCT+TAU compared to TAU. This neural change was associated with reduced trait rumination and depressive symptoms, suggesting that it may play a role in reducing the persistence and “stickiness” of ruminative thought patterns (van der Velden, 2025). All these different scientific approaches are paving the way to unpacking the neural reorganisation accompanying MBCT treatment from different perspectives using increasingly sophisticated methods. To date, no study has investigated changes in hierarchical brain processing, a key organisational principle thought to enable efficient distributed spatiotemporal computations necessary for survival and optimal functioning (Deco et al., 2021). Moving beyond descriptive neural studies toward uncovering the underlying brain hierarchical organisation could help explain how MBCT promotes psychological flexibility and reduced vulnerability to depression.

The ‘Thermodynamics of the Mind’ is a promising framework for studying hierarchical brain dynamics and its underlying mechanisms (Kringelbach et al., 2024). Evidence from neurology (e.g., Alzheimer’s (Cruzat, 2023), stroke (Idesis et al., 2023), coma (García Guzman et al., 2023)) and neuropsychiatry (Socoro-Garrigosa et al., 2025; Vohyzek) highlights the framework’s clinical promise, including findings showing different treatment response to pharmacological interventions in MDD (Deco, 2024). This last investigation implemented a measure of directedness on whole-brain models. The method was inspired by work in ecology which illustrates the hierarchical relationships between components in a food web by assigning a trophic level (i.e., position of node in the hierarchy) to each node in a directed graph. This allows to capture the global directedness (i.e., directionality of information flow), cycle structure, and stability (MacKay et al., 2020). In this way, equal trophic levels reflect a flat global hierarchy, low directedness, and high recurrence, while a multi-layered network represents a high global hierarchy, high directedness, and low recurrence. As such, this method is key to uncover functional brain hierarchy reorganisation underlying treatment-related changes following mindfulness training for MDD.

In this novel secondary analysis, we explored how MBCT affects brain hierarchy in patients with MDD in our randomised controlled trial using the approach described by (Deco, 2024). We built whole-brain models and obtained a generative effective connectivity (GEC) matrix of each patient in each treatment, session, and scan condition. Then, we quantified brain hierarchy by measuring the global directedness and trophic levels of each area in the GEC. To our knowledge, this is the first study quantifying brain hierarchical changes in MBCT for MDD and complements previous investigations providing insights into our understanding of the underlying brain changes towards recovery. Overall, our analysis may create new opportunities for the identification of individuals responding to treatment, brain markers, and new experimental designs in the field of MDD.

## Materials and Methods

### – Study design

The design of the trial, previously published (van der Velden, 2023), consisted of a single-blind, randomised controlled trial of Mindfulness-Based Cognitive Therapy (MBCT) in addition to treatment as usual (TAU) (MBCT+TAU), or TAU alone, to investigate the changes in neural mechanisms and psychological processes (**Figure 1A-B**). The study was approved by the Regional Ethics Committee of Central Denmark (ID: 1-10-72-259-16: 66534). It was registered at ClinicalTrials.gov (identifier: NCT03353493) and the Danish Data Protection Agency (2016-051-000001). For details on participant (n=80) recruitment please see **Supplementary Information**.

**Figure 1.**
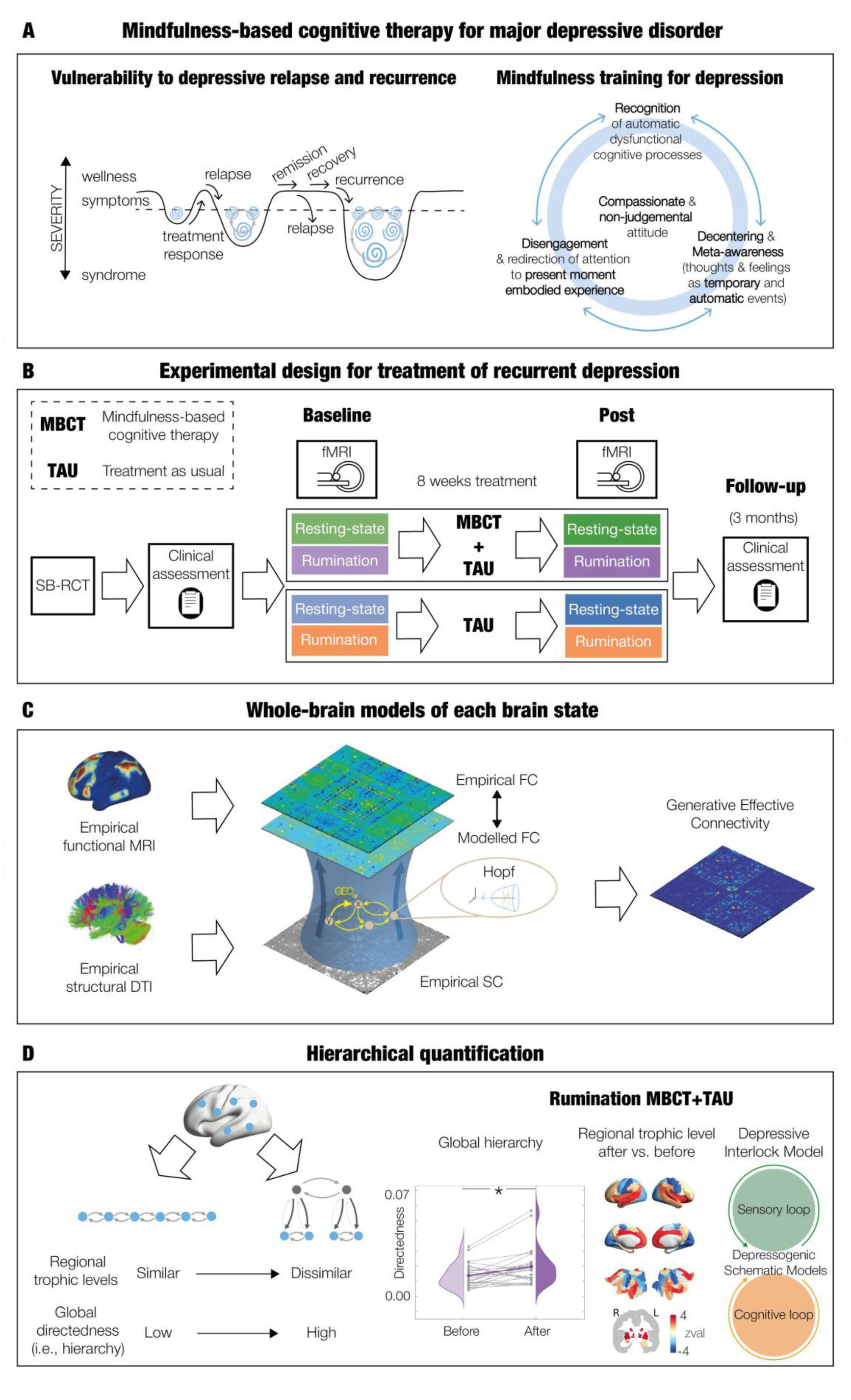
Methodology for assessing changes in brain hierarchy in MBCT treatment for depression. **A.** The left plot schematises the progression of major depressive disorder ( MDD) from acute phase to response to treatment, remission and recovery, and the risk for relapse and recurrence. The x-axis represents the phases of treatment and the y-axis the symptom severity. The curving valleys symbolise episodes of depression over time, with the depth corresponding to the severity of symptoms and the spirals echoing rumination. Response is when symptoms start to improve; remission corresponds to the period in which symptoms are largely gone but there is vulnerability to relapse; and relapse and recurrence are the returning to an episode during/shortly after treatment is completed, or after recovery, respectively; and recovery is identified after sustained remission during longer periods of time. Schematic adapted from (Segal, 2013). The right plot illustrates the key processes of mindfulness-based cognitive therapy program consisting of recognition of automatic dysfunctional cognitive processes, decentering and meta-awareness (thoughts and feelings as temporary and automatic events), and disengagement and redirection of attention to present moment embodied experience, all with a compassionate and non-judgemental attitude (Segal, 2013). **B.** Single blind randomised controlled trial (SB-RCT) study design of Mindfulness-Based Cognitive Therapy (MBCT) in addition to treatment as usual (TAU) (MBCT+TAU), or TAU alone. It includes randomisation, clinical assessment and pre– and post fMRI collection. **C.** Whole-brain models are optimised and fitted to the neuroimaging data of each participant, creating Generative Effective Connectivity (GEC) matrices. **D**. Inspired by previous work in ecology and extended to general directed networks, in the GEC matrices each brain region is assigned a trophic level (i.e., hierarchical position), and the global directedness (i.e., trophic coherence) can be calculated. A flat hierarchy is governed by similar trophic levels and low directedness. In contrast, a strong hierarchy presents more layers and diversity in trophic levels, associated with high directedness. Figure adapted from (Deco, 2024) under Creative Commons Attribution License 4.0 (CC BY).

### – Intervention

The two groups of participants consisted of either MBCT+TAU, or TAU alone. Mindfulness-based cognitive therapy is an 8-week group program aimed to prevent relapse or recurrence to depression (Segal, 2013). It combines psychoeducation elements from cognitive behavioural therapy for depression, and systematic training in mindfulness meditation from the mindfulness-based stress reduction program. The training was delivered according to the treatment manual (van Aalderen et al., 2015), in university settings, by experienced therapists. Therapists were highly experienced MBCT teachers (≥7 years) based on international guidelines (Crane & Kuyken, 2019; Kuyken, 2012) and held the highest competency rating (Fresco et al., 2007). Overall, the therapy consisted of a pre-class interview, weekly classes of 2.25 hours each, daily homework during the 8 weeks period of treatment, a whole practice day near the 6^th^ session, and 4 booster sessions offered every 3 months after the program.

The TAU generally consists of antidepressant medication (ADM) and psychological therapy. Here, we restricted it to either no medication, or a stable dose of antidepressant medication. Participants on medication were encouraged to adhere to their regimen throughout the whole trial and asked to report any changes, without affecting their participation.

Participants were assessed with questionnaires and MRI-scans before and after treatment and depressive symptoms were measured again at three months follow up. They also provided qualitative feedback following the intervention open ended survey responses. Engagement with the treatment and practice was also recorded for the MBCT+TAU group. Mean attendance was above 7 (out of 8 sessions) and mean practice was above 3 days (per week).

### – Clinical measures and psychological processes

The following self-reported measures were assessed: depressive symptoms using the Quick Inventory of Depressive Symptomatology-Self Report (QIDS) (Rush et al., 2003), perceived stress using the Perceived Stress Scale (PSS) (Cohen et al., 1983); interoceptive awareness using the subscales of noticing (NO), emotional awareness (EA), body listening (BL), attention regulation (AR), trusting (TR), and not-distracting (ND) of the Multidimensional Assessment of Interoceptive Awareness (MAIA) (Mehling et al., 2012), decentering using the Experiences Questionnaire – decentering factor (Fresco et al., 2007), mindfulness skills using the Five Factor Mindfulness Questionnaire – short version (FFMQ) (Baer et al., 2008; Tran et al., 2013) and trait rumination using the Rumination Response Scale (RRS) (Langenecker et al., 2024; Roelofs et al., 2006).

### – fMRI paradigm

The fMRI paradigm included an initial structural scan followed by 4 consecutive and separate functional connectivity scans of 5 minutes each at different conditions. The order was of resting-state, instructed mindfulness state, resting-state, and instructed rumination state. Before each scan, the research team instructed the participants on the nature of the scan condition and, for ethical reasons, highlighted the voluntary aspect. Participants could opt out of rumination if they felt it was too stressful. After each scan condition, an experience sampling in a computer screen inside the scanner was done assessing affective, cognitive and somatic experiences, adapted from (Smallwood et al., 2016). It consisted of a visual analog scale (VAS) and a cursor that moved with a trackball to indicate the degree of agreement from 0-100%. There was also a brief follow-up talk with a clinically trained member to make sure participants were okay.

In this work we focus on the changes before and after treatment in the first resting-state and the rumination induction. The former because it is related to the general vulnerability of depression and, given it is measured first, we avoid potential confounding carry-over effects from the rest of the scan conditions. And the latter since MBCT targets depressive rumination. We validated the effectiveness in rumination with VAS scores (**Supplementary Figure S1**). For more details on MRI paradigm and rationale please see **Supplementary Information**.

### – Functional brain hierarchy metrics

Following (Deco, 2024), we built a whole-brain model for each patient using Stuart-Landau oscillators to represent the local dynamics of each brain region (90 brain areas using the Automated Anatomical Labelling atlas (Tzourio-Mazoyer et al., 2002) (**Figure 1C**). This corresponds to the normal form of a supercritical Hopf bifurcation, capable of describing transitions from asynchronous noise to oscillations. From the whole-brain models, we created generative effective connectivity (GEC) matrices in which existing anatomical connectivity strength is adapted iteratively until best fit to empirical fMRI data. For a full mathematical description of the Hopf model, its linearization and optimisation, please see **Supplementary Information**. Then, we implemented brain hierarchy measures of trophic levels and directedness in directed networks, to the individual GEC matrices (**Figure 1D**). We computed the trophic level for each brain region and the global directedness (i.e., trophic coherence) of the whole-brain.

#### Trophic levels

The GEC matrix defines a graph with *N* nodes, connected by weighted edges, determined by *C*. For each node *n*, the in-weight 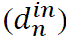 and the out-weight 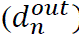 are calculated by summing the *m* columns and rows of the matrix, respectively, as follows:

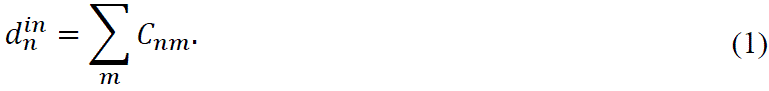

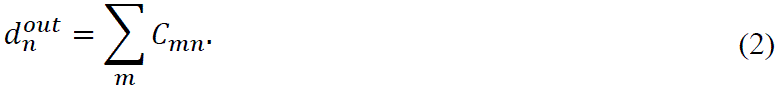

The total weight of a node *u_n_* is defined by

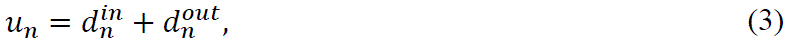

and the imbalance *v_n_* as the difference between the flow into and out of a node:

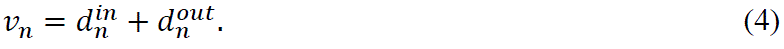

Furthermore, the weighted graph-Laplacian operator ∧ on vector **h** is

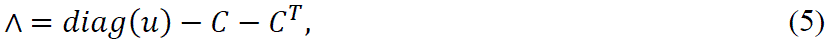

and the solution **h** of the following linear system of equations can be obtained:

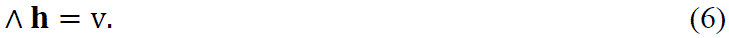

The trophic level of each brain area corresponds to each component of **h**. As a note, the operator ∧ is symmetric and the asymmetry comes from the imbalance vector v.

#### Directedness (trophic coherence) of a network

The global directionality (i.e., directedness or trophic coherence) is calculated with the following equation:

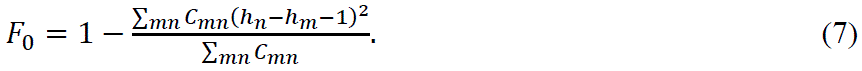

Maximally coherence corresponds to *F*_0_ = 1, whereas incoherence is regarded when *F*_0_ = 0.

### – Statistical Analysis

We performed all statistical analysis using Wilcoxon sign-rank and rank-sum and reported the p-values. In the analysis of brain markers at a participant level, we implemented permutation-based statistics (permutations=1000, significance threshold=0.05). In the analysis of brain markers at a node-level, we reported the z-value (standardised test statistic). We corrected for multiple comparisons, when applicable, implementing False Discovery Rate (FDR) method (Hochberg & Benjamini, 1990).

### – Comparisons of brain measures with psychological processes and clinical outcomes

We evaluated the relation between brain measures and psychological processes and clinical outcomes using Spearman and partial Spearman correlations. We performed mediation analysis via bootstrap test (10,000 permutations) using a mediation toolbox (Wager et al., 2008).

## Results

We studied the spatiotemporal hierarchical organisation of brain dynamics in our randomised controlled trial of MBCT+TAU, or TAU, as treatment for MDD (**Figure 1A-B**). First, we built whole-brain models integrating underlying anatomical structure with functional brain dynamics and obtained a GEC matrix for each patient of each treatment (MBCT+TAU and TAU), scan condition (resting-state and rumination induction state) and session (before and after). We fitted the functional connectivity and its shifted version to capture the asymmetry in information flow (**Figure 1C**). Then, we calculated the regional trophic levels in each GEC and global directedness (**Figure 1D**).

Participant demographics and baseline psychiatric variables and questionnaire scores were similar between the treatments (**Supplementary Table S1**). The size of the cohort analysed was N=39, N=26, N=26 and N=21 for MBCT+TAU resting-state, TAU resting-state, MBCT+TAU rumination, and TAU rumination, respectively (Consort diagram in **Supplementary Figure S2**). Fewer participants performed rumination compared to resting-state as, for ethical motives, the condition was voluntary. The participants not undertaking rumination (∼28%, n=18 from whole sample of n=65) had similar baseline characteristics than the participants performing resting-state and rumination, or rumination only, except for age and depressive symptoms, with more individuals in the severely depressed range and less below the symptomatic threshold (**Supplementary Figure S3A**). After MBCT treatment, both those who participated in the rumination condition and those who did not, experienced a reduction in depressive symptoms (**Supplementary Figure S3B**).

### – Global directedness in brain states

Results showed a significant increase in global directedness during rumination in MBCT+TAU (p < 0.05), which remained significant after excluding the two subjects with consistent extreme positive values across sessions (**Supplementary Figure S4**). No significance was found during rumination in TAU nor during resting-state in either intervention group (MBCT+TAU or TAU) (**Figure 2A**). Baseline global directedness did not differ between treatments or conditions whereas in the post-session the global directedness in rumination was significantly higher than resting-state within MBCT+TAU (p < 0.001), an effect not observed in TAU (**Figure 2B**). Analysis of the change (after-before) revealed a larger significant increase in global directedness in MBCT+TAU compared to TAU during rumination (p < 0.05), with no intervention group differences in resting-state. Furthermore, there was a significant condition-specific change in MBCT+TAU, stronger in rumination compared to resting-state (p < 0.01), and not in TAU (**Figure 2C**).

**Figure 2.**
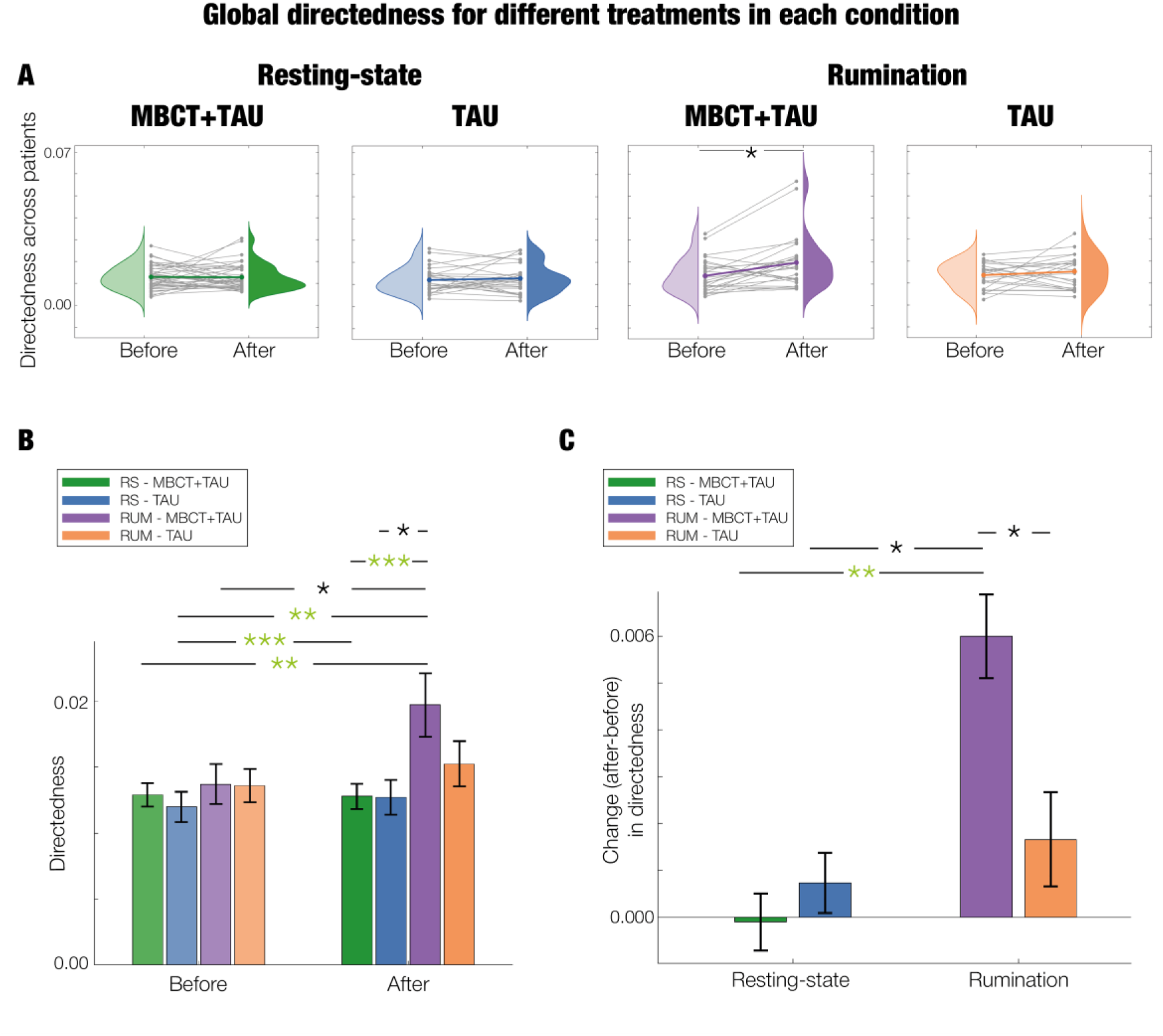
Global hierarchical reconfiguration with treatment. In all plots, significance is represented with asterisks (*, p < 0.05; **, p < 0.01; ***, p < 0.001), in green if they survive correction for multiple comparisons. **A.** At a global level, significant differences in directedness between the before and after treatment are found only in MBCT+TAU rumination. Grey lines represent the trajectory of each individual whereas the coloured line represents their averaged trajectory within each intervention arm and condition. **B.** Directedness at baseline and post-treatment. Bar plots showing the mean and standard error of the mean for all treatments (MBCT+TAU and TAU), condition (resting-state and rumination), and session (before and after). **C.** Comparing the change (after-before) in directedness for MBCT+TAU, and TAU, in resting-state and rumination. Barplots show the mean of the change and error bars show the standard error of the mean of the change. A linear mixed model confirms the differential effect of treatment*condition.

We also fit a linear mixed-effects model predicting global directedness from treatment (1: MBCT+TAU, 2: TAU), condition (1: resting-state, 2: rumination), and their interaction, with a random intercept for participant ID. The model revealed no significant effects on treatment (β = 0.006, SE = 0.004, p = 0.110), and significant effects on condition (β = 0.012, SE = 0.004, p = 0.002) and the treatment × condition interaction (β = –0.005, SE = 0.002, p = 0.029). Significance remained when controlling for covariates of age, sex and ADM – treatment (β = 0.006, SE = 0.004, p = 0.114), condition (β = 0.012, SE = 0.004, p = 0.001), and treatment × condition interaction (β = –0.005, SE = 0.002, p = 0.029). Overall, the effects of MBCT+TAU and TAU on global directedness were condition-dependent, with a larger resting-state to rumination change increase in MBCT+TAU compared to TAU.

### – Regional differences in trophic levels

We then computed the trophic level of each brain area to uncover the regional reconfigurations following each treatment and scan condition. Brain renders of the regional trophic levels averaged across individuals can be seen in **Figure 3A**, and the brain areas with highest and lowest values are listed in **Supplementary Tables S2-5**. In all cases, brain areas with highest trophic levels had an increased imbalance (i.e., higher in-flow compared to out-flow of effective connections) (**Supplementary Figure S5**). Furthermore, statistics (z-value and significance) between the pre– and post– trophic levels within each treatment and scan condition are shown in **Figure 3B** and listed in **Supplementary Tables S6-9**. Lastly, regional differences of the changes (after-before) between MBCT+TAU and TAU within each condition (resting-state and rumination), and between resting-state and rumination within each treatment (MBCT+TAU and TAU) and session (before and after) can be observed in **Supplementary Figure S6**. Overall, highest regional changes and significance corresponded to MBCT+TAU during rumination, whereas minimal changes were observed for the rest of the groups.

**Figure 3.**
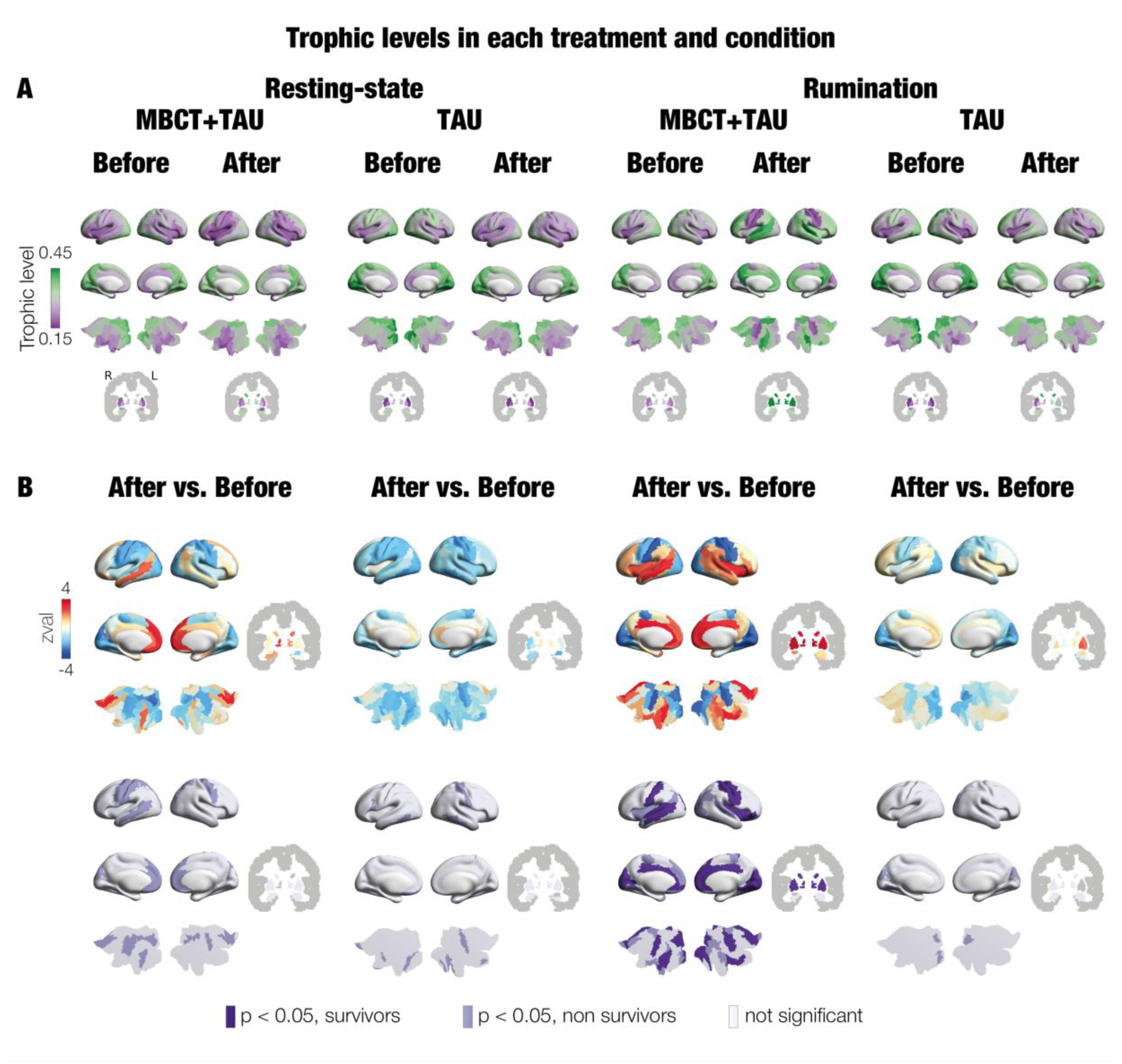
Regional trophic level reconfiguration with treatment. **A.** At a regional level, the average trophic levels across patients are rendered on brain maps for each condition (resting-state and rumination), treatment (MBCT+TAU and TAU) and session (before and after). **B**. Statistical differences in trophic levels between after and before each condition and treatment. Top row shows z-values with positive/negative values, in red/blue, corresponding to higher/lower trophic levels after treatment compared to baseline. Second row shows p-values, in dark purple significant brain areas surviving correction by multiple comparisons, in lilac significant brain areas not surviving correction by multiple comparisons, and in white non-significant brain areas. Subcortical regions are shown on slices in Montreal Neurological Institute (MNI) space (coronal axis y= –6 mm).

The trophic framework quantifies the magnitude in asymmetric magnitude of interactions captured in the GEC matrices fitted to the empirical data. We applied the INSIDEOUT framework (Deco et al., 2022) to evaluate the asymmetry in information flow in the empirical fMRI signals, namely the irreversibility (**Supplementary Methods and Supplementary Figure S7**). This method has shown to be robust and sensitive in distinguishing different brain states (García Guzman et al., 2023; Kringelbach et al., 2023). Results were consistent with our previous model-based analysis (**Supplementary Figure S8 and S9**).

### – Rumination MBCT+TAU: response to treatment

Given that rumination showed the greatest sensitivity at both global and regional levels in the MBCT+TAU group, we further examined brain area reconfiguration and its association with clinical and behavioural measures specifically for this scan condition and intervention group.

#### Regional reconfiguration

First, we examined trophic level changes associated with Yeo resting-state networks (Yeo et al., 2011). After treatment, regions with highest contribution to the DMN and subcortical areas increased and started having highest trophic levels, whereas the regions highly associated with the visual network (VIS) and some of the SOM decreased and showed lowest trophic levels. Furthermore, the regions related mainly to the SOM and subcortical areas increased their spread along the trophic level distribution (**Supplementary Figure S10**).

Secondly, we computed the trophic levels for three different symptomatic trajectory groups defined by changes in depressive symptoms, individuals who remained asymptomatic across sessions, transitioned from symptomatic to asymptomatic, or remained symptomatic from pre– to post-sessions (**Supplementary Figure S11 and Supplementary Tables S10-12**). The symptomatic-to-asymptomatic group after treatment exhibited the largest distance between the upper and lower ends of the trophic level distribution and most closely resembled the effects observed in the full sample of rumination MBCT+TAU (**Figure 3A**).

Thirdly, we examined the distribution of trophic levels, finding it was skewed towards the positive tail across sessions (**Supplementary Figure S12A**). This skewness was more pronounced post-treatment and led the higher dispersion in trophic levels. We quantified this effect by calculating the breadth of hierarchy, defined as the distance between the trophic levels in the extremes of the distribution. Results showed significant increases after treatment consistent with the changes in global directedness (**Supplementary Figure S12B**).

#### Relation with clinical and behavioural outcomes

Subsequently, we evaluated the relation between brain changes with clinical and behavioural outcomes during rumination in the MBCT+TAU group. We implemented Spearman and Partial Spearman correlation controlling for age, sex, ADM and baseline depressive symptoms to mitigate confounding effects and ensure robust association estimates. As brain marker, we used the breadth of hierarchy calculated as the distance between regions above and below 1 standard deviation given it resulted in the highest significance across sessions (**Supplementary Figure S12B**). As for the clinical and behavioural outcomes, we selected the scores presented in **Supplementary Table S1**. For reference and descriptive purposes, we report results for the TAU group, and the whole sample (MBCT+TAU and TAU), in the **Supplementary Information**. Given the exploratory nature of this post hoc analysis, no correction for multiple comparisons was applied.

First, we studied the association between the change (after-before) in clinical and behavioural outcomes with the change (after-before) in breadth of hierarchy (**Supplementary Table S13**). For the Spearman correlation, it was significantly negative for perceived stress (rho = –0.505, p = 0.010) and rumination (rho = –0.471, p = 0.017), and positive for mindfulness (rho = 0.448, p = 0.025), decentering (rho = 0.645, p < 0.001) and the attention regulation (AR) subscale of interoceptive awareness (rho = 0.609, p = 0.001). These were still significant with Partial Spearman correlation, negative for perceived stress (rho = –0.562, p = 0.008) and rumination (rho = –0.509, p = 0.018), and positive for mindfulness (rho = 0.529, p = 0.014), decentering (rho = 0.761, p < 0.001) and AR (rho = 0.660, p = 0.001), in addition to new significant correlations, negative with depressive symptoms post-treatment (rho = –0.481, p = 0.023) (**Figure 4A**), and positive with the body listening (BL) subscale of interoceptive awareness (rho = 0.471, p = 0.031).

**Figure 4.**
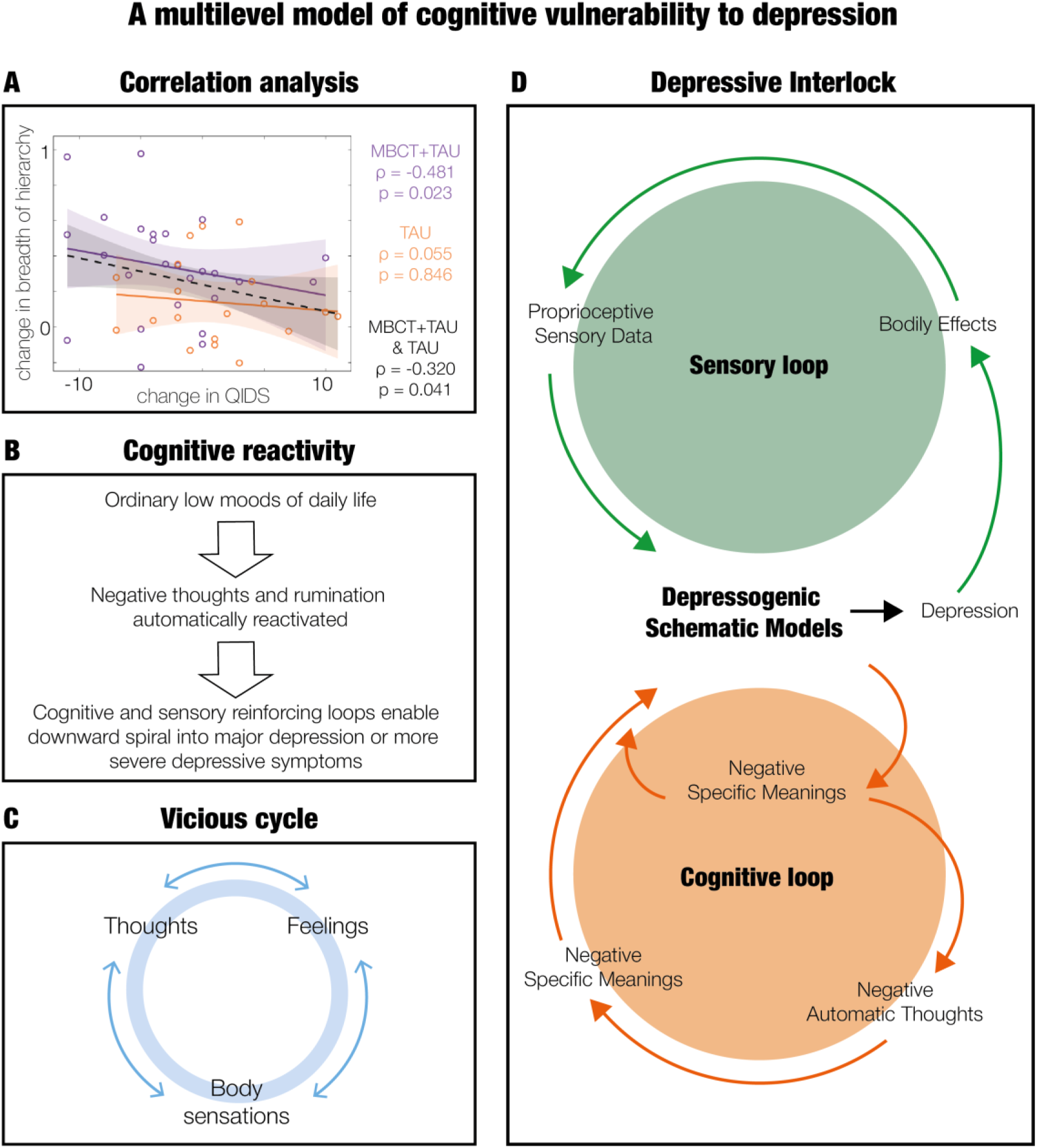
A multilevel model of cognitive vulnerability to depression. MBCT addresses vulnerability to depression through prevention (**B** and **C**) and maintenance (**D**) models. **A.** Correlation between change (after-before) in breadth of hierarchy and change (after-before) in depressive symptoms post treatment. The plot shows the correlation for the MBCT+TAU group in violet, the TAU group in orange, and both groups together (MBCT+TAU and TAU) in black. Each circle represents a participant and the lines the trends in change. Significance is found for the MBCT+TAU group, and when combining the two intervention arms, with increased breadth of hierarchy related with improvements in depressive symptoms. **B.** Everyday low moods in people who had depression in the past can quickly reactivate old patterns of negative thinking and bodily feelings. The cognitive and sensory reinforcing loops can drive a downward spiral into major depression or more severe symptoms (Teasdale, 1988). **C.** Once cognitive reactivity has been triggered, thoughts, feelings and bodily sensations keep each other ongoing in a self-reinforcing vicious cycle (Segal, 2013). **D.** The internal maintenance of depression can be conceptualised as a self-perpetuating ‘Depressive Interlock’ processing configuration in which cognitive and sensory loops interact to sustain depressive states once they are activated. At the centre of this configuration are the depressogenic schematic models, representing mental high-level regularities in the world, body and mind. In the cognitive loop (orange), the depressogenic schematic models generate negative meanings (i.e., negative evaluations of one-self, the future, interpersonal relations). These meanings are experienced as streams of negative automatic thoughts. These further reinforce and elaborate on the negative meanings, in turn feeding and strengthening the models. In the sensory loop (green), depressive schematic models produce depressive emotional reactions that include bodily effects (e.g., sad, frowning expression, tearful) which lead to body-state information. This contributes, together with the meaning-derived elements from the cognitive loop, to perpetuating the ‘depressive interlock’ and in consequence maintaining the production of depression. Within this framework, depression persists as long as the depressogenic schematic models continue to be produced and treatments should aim to prevent the re-establishment of depressive interlock configurations. Schematic adapted from (Teasdale et al., 1995).

Second, we studied whether baseline clinical and behavioural scores were related with the change in breadth of hierarchy (**Supplementary Table S14**). For the Spearman correlation, it was significantly negative for baseline decentering (rho = –0.497, p = 0.011), AR (rho = –0.493, p = 0.012) and BL (rho = –0.445, p = 0.026). These were still significant with Partial Spearman correlation for decentering (rho = –0.588, p = 0.005), and AR (rho = –0.436, p = 0.048) except for BL.

Third, we performed a mediation analysis to evaluate if treatment effects on changes in clinical and behavioural outcomes were mediated by brain marker, and vice versa (**Supplementary Tables S15 and S16**). For the non-adjusted analysis, treatment influenced changes in perceived stress, mindfulness, rumination, decentering, the emotional awareness (EA) subscale of interoceptive awareness and AR, mediated by the change in breadth of hierarchy. The reciprocal models showed treatment influenced change in breadth of hierarchy via all previous scores except for rumination. The covariate-adjusted mediation analyses showed treatment influenced change in perceived stress, rumination and EA, mediated by the change in breadth of hierarchy, and the reciprocal models showed treatment influenced change in breadth of hierarchy via perceived stress.

In order to examine brain changes of response without prior assumptions, we employed an unsupervised clustering technique to uncover changes in brain trophic level patterns associated with how much participants improved clinically, relative to their baseline depressive symptoms. Results showed a regional configuration of trophic levels with opposite directions of change (and stronger changes for the positive areas), associated with improvements in depression symptoms (**Supplementary Figure S13**).

## Discussion

In this novel secondary analysis, we explored the hierarchy in brain dynamics of patients with major depressive disorder (MDD) receiving mindfulness-based cognitive therapy (MBCT) in addition to treatment as usual (TAU), or TAU alone, during resting-state and rumination from our randomised controlled trial (van der Velden, 2023) (**Figure 1**). We found that global directedness in MBCT+TAU compared to TAU increased during rumination, with no changes during resting-state. In addition, increased regional breadth of hierarchy during rumination was related to improvements in clinical and behavioural outcomes following MBCT+TAU. Overall, this work builds upon our previous work implementing both empirical static (van der Velden, 2023) and dynamic (van der Velden, 2025) analysis, moving beyond descriptive neural studies toward uncovering the potential underlying brain processes within hierarchical brain dynamics. To our knowledge, this is the first study uncovering how hierarchical brain processes change during rumination in response to MBCT treatment for MDD.

At a global level, we found a significant increase in directedness during rumination in MBCT+TAU and not TAU, and no differences during resting-state for either group (**Figure 2**). The increased directedness in rumination with MBCT+TAU was indicative of more hierarchical global brain dynamics and less recurrence. This was accompanied with a regional reconfiguration after treatment, with brain regions belonging mainly to the visual and somatomotor network showing lowest trophic levels, and areas corresponding mainly to the default mode network and subcortical areas showing highest trophic levels (**Figure 3**). This strong opposite direction of change was associated with a prototype of brain response to positive clinical improvements (**Supplementary Figure S13**). Interestingly, the change seems to follow the well-known cortical gradient-like organisation with unimodal regions (i.e., low-order cognitive networks) in one end, and transmodal association regions (i.e., high-order cognitive networks integrating different sources of information) in the other end. This spatial arrangement has been identified as key for human cortical organisation along a global gradient, reflecting the range of functions from direct perception and action, to processing and integration of abstract information (Bernhardt et al., 2022; Huntenburg et al., 2018; Margulies et al., 2016). A past study found patients with MDD exhibited narrower range and smaller variance of this principal gradient, suggesting that the distance along the cortical gradient is key for ensuring a processing route from concrete stimuli to integration of abstract cognition avoiding interferences of input noise (Xia et al., 2022).

Furthermore, several of the regions that showed the largest changes are consistent with our previous publications in this dataset, including the anterior cingulate cortex, insula and lingual gyrus (van der Velden, 2023), of which the first two are commonly implicated in mindfulness and psychotherapy related effects, as well as in depression research (Young et al., 2018) (Marwood et al., 2018) (Godlewska et al., 2018). In addition, other regions showing increased changes were somatomotor areas such as postcentral gyrus and temporal superior gyrus (van der Velden, 2025), which have been linked to depression (Fan & Zhang, 2025; Kandilarova et al., 2019).

To further understand the regional changes during rumination following MBCT+TAU, we quantified the novel measure breadth of hierarchy and evaluated its association with clinical and behavioural outcomes. We first defined breadth of hierarchy as the distance between the regional trophic levels in the extremes of the hierarchical distribution, showing significant increases in line with the global positive changes in directedness (**Supplementary Figure S12**). We then applied Partial Spearman correlation corrected for age, sex, ADM and baseline QIDS. Results showed that changes in regional breadth of hierarchy during rumination MBCT+TAU were associated with improvements in depressive symptoms (**Figure 4A**), perceived stress, rumination, mindfulness, decentering, active listening to the body for insight and ability to sustain and control attention to body sensations (i.e., body listening and attention regulation subscales of interoceptive awareness, respectively) (**Supplementary Table S13**). In addition, individuals who at baseline reported a lower ability to be able to decenter from ruminative negative thoughts and to sustain and control attention to body sensation experienced greater change in breadth of hierarchy after MBCT treatment (**Supplementary Table S14**). Lastly, a mediation analysis, corrected by the aforementioned covariates, further indicated that treatment-related changes in perceived stress and rumination were mediated by changes in breadth of hierarchy, with reciprocal mediation persisting for perceived stress (**Supplementary Tables S15 and S16**). This suggests a dynamic interplay between neural and clinical and behavioural changes. Overall, our findings position regional breadth of hierarchy as a possible MBCT+TAU treatment-sensitive brain marker that relates to both clinical and behavioural improvements as well as baseline profiles during rumination.

We suggest our findings on rumination with MBCT+TAU could be interpreted, with caution, with well-established theories of psychological vulnerability to depression. The cognitive reactivity framework (**Figure 4B**) proposes that mild negative affect can trigger habitual ruminative vicious cycles (**Figure 4C**), which may escalate into depressive episodes when such patterns become increasingly rigid and self-reinforcing (Segal, 2013). Depressive symptoms generally tend to worsen across episodes (reflected in the increasing depth of “valleys” in **Figure 1A**), increasing the probability of future episodes (van der Velden, 2015), potentially reaching a point of helplessness and powerlessness closely linked to elevated suicide risk (Chesin et al., 2016). A more elaborated account is provided by the ‘Depressive Interlock’ processing configuration (Teasdale et al., 1995). This model describes how two interacting self-reinforcing loops – cognitive self-referential and sensory-affective processing – jointly can perpetuate a depressive state by regenerating depressogenic schematic models (i.e., mental high-level regularities in the world, body and mind). As such, the depressed state can resolve if the re-establishment of depressive interlock configuration is prevented (detailed description in **Figure 4D**). Within this theoretical context, the global and regional brain changes observed following MBCT+TAU during rumination could reflect a shift away from recurrent self-referential and sensory-affective loops towards a more differentiated, hierarchical, and directional information flow. Higher-order regions could thereby better integrate and regulate emotional and bodily signals while allowing cognitive signals to propagate more effectively toward sensorimotor areas. Without overestimating the implications of our results, such brain changes may provide a mechanistic account of core MBCT processes as a means of interrupting automatic entrenched cycles between cognitive and sensory-affective loops described in the ‘Depressive Interlock’ (Teasdale et al., 1995). This way, the prevalence of mood-induced negative thoughts and ruminative tendencies after MBCT therapy might still exist, but the perpetuating tendencies and “stickiness” of the ruminative thoughts may change. Future work could focus on further unpacking the role of brain hierarchy in depression by examining differences between patients with MDD and healthy controls. Furthermore, longer follow-up periods would allow testing whether rumination-evoked changes in hierarchical brain organisation can predict subsequent relapse or recurrence.

Other important insights from this study include neural changes observed following MBCT+TAU and not TAU (**Figure 2A**), supporting the role of MBCT as an effective intervention for modulating neurocognitive functioning. In addition, rumination and resting-state were significantly different after MBCT+TAU and not at baseline (**Figure 2B**). The non-significant brain dynamics at baseline suggests rumination and resting-state are less distinguishable in MDD. And, the fact that after treatment they were significantly different could suggest participants learn to adapt to the different cognitive-emotional demands following mindfulness training. Furthermore, their different direction of change emphasises the importance of understanding treatment changes can be state dependent, as is the case of stimulation experiments for example (Bradley et al., 2022). Moreover, given significance was only found during rumination, as in previous studies in the same dataset (van der Velden, 2023; van der Velden, 2025), suggests it is a sensitive context for characterising depression at a neural level, and we speculate – while avoiding reverse inference – that depression may reflect an inability to disengage from ruminative cycles (Holtzheimer & Mayberg, 2011).

Our work presents several limitations. First, the choice of TAU as a control group, although it allows for external validity as a real-life application, lacks specificity as a control condition. Given the absence of an evidence-based active control group, we cannot infer whether changes are specific to MBCT or similar efficacy levels can be obtained from other psychotherapeutic treatments. Another limitation is that, for ethical reasons, participation in the rumination condition was voluntary. Participants who opted out, while spanning asymptomatic/mild to severe depression as participants performing rumination, had higher depressive symptoms at baseline. In consequence, findings in rumination are applicable only to this subsample. Finally, we acknowledge that despite our efforts in trying to understand the underlying brain changes using theoretical methods in the field of neuroscience, there is still an explanatory gap between neural processes and subjective experience. And, we are aware of the loss of ecological validity in translating real-life MBCT effects into laboratory settings.

To conclude, this novel secondary analysis provides the first investigation of functional brain hierarchy in MBCT for major depressive disorder. During rumination following MBCT+TAU, we observed increases in global directedness and regional breadth of hierarchy related with behavioural and clinical improvements. We suggest a possible association with psychological theories describing cognitive vulnerability to depression, reflecting a shift away from self-reinforcing negative metal loops towards more differentiated cognitive and bodily thinking processes, interrupting ruminative thinking patterns. Overall, these findings contribute to a more integrated account of MBCT related brain and psychological changes, and underscore hierarchical brain dynamics as a promising framework for studying depression treatments.

## Data availability

The data can be requested to the corresponding authors upon reasonable request.

## Code availability

FSL software is available at https://fsl.fmrib.ox.ac.uk/fsl/fslwiki/FslInstallation. Whole-brain models and analysis was performed in MATLAB R2017b, R2022a and R2024a software from MathWorks (Natick, MA, USA). The code is available from the corresponding authors upon request.

## Acknowledgements

This research was funded by a Carlsberg Foundation Internalization Fellowship (CF21_0645) to A.M. P.D. is supported by the Department of Research and Universities of the Government of Catalonia, Agency for Management of University and Research Grants (AGAUR), FI-SDUR programme (Grant No. 2022 FISDU 00229). H.G.R. is supported by grants from the Hersenstchting, ZonMW, Parkinson Foundation, and Radboudumc. He obtained unrestricted educational grants from J&J and served as a speaker for J&J, Lundbeck, E-wise, Prelum and Benecke. M.L.K. is supported by the Centre for Eudaimonia and Human Flourishing (funded by the Pettit and Carlsberg Foundations) and Center for Music in the Brain (funded by the Danish National Research Foundation, DNRF117). J.V. is supported by the EU funded Project NEurological MEchanismS of Injury, and Sleep-like cellular dynamics (NEMESIS; ref. 101071900) funded by the EU ERC Synergy Horizon Europe. G.D. is supported by Grant PID2022-136216NB-I00 funded by MICIU/AEI/10.13039/501100011033 and by “ERDF A way of making Europe”, “ERDF, EU”, Project NEurological MEchanismS of Injury, and Sleep-like cellular dynamics (NEMESIS) (ref. 101071900) funded by the EU ERC Synergy Horizon Europe, AGAUR research support grant (ref. 2021 SGR 00917) funded by the Department of Research and Universities of the Generalitat of Catalunya, and Grant PID2024-155136NI-I00 financed by MICIU/AEI/10.13039/501100011033/ and by “ERDF A way of making Europe”, ERDF, EU.

