## Supplementary Information for "Changes in hierarchical brain dynamics of rumination following mindfulness-based cognitive therapy for depression"

### Materials and Methods

#### - Participants

Participants (n=80) were recruited from psychiatric units and general practices in Central Jutland region, Denmark. Inclusion criteria were: diagnosis of recurrent major depressive disorder (with or without a current episode) using Structured Clinical Interview for DSM-IV-TR (Gorgens, 2011); 3 or more previous major depressive episodes; age of 18 or older; if on antidepressant medication, a stable dose for a minimum of 8 weeks of selective serotonin re-uptake inhibitor, or serotonin and norepinephrine reuptake inhibitor. The DSM defines a depressive episode by at least two weeks of depressed mood (i.e., sadness) and loss of interest, together with four or more additional symptoms such as feelings of worthlessness, impaired concentration, or fatigue (Association, 2013).

Exclusion criteria were: current severe major depressive episode using Beck Depression Inventory-II > 28 (Beck, 1996); history of schizophrenia, schizoaffective disorder, bipolar disorder; current severe substance abuse, organic mental disorder; current/past psychosis, pervasive development delay, persistent antisocial behaviour, persistent self-injury requiring clinical therapy or management; concurrent formal psychotherapy; previous completion of MBCT/mindfulness-based stress reduction training; exhaustive meditation experience (i.e., regular practice, retreats); or antipsychotic medication and benzodiazepines. Written informed consent was given by all participants.

#### - Randomisation and blinding

An independent researcher randomly allocated participants to receive either an 8-week MBCT+TAU training, or adhere to TAU, in a 5:3 ratio. Randomisation was performed using a computerised system and stratified based on antidepressant use and participants' symptoms using Beck Depression Inventory-II (asymptomatic for =<13 and symptomatic for >=14) (Beck, 1996). This way, to ensure the distribution of patients taking or not antidepressant medication, and proportion of symptomatic and asymptomatic patients, was balanced across the two groups. Furthermore, sociodemographic and clinical characteristics were also balanced between the groups. Participants, therapist and trial coordinator were masked to treatment allocation at baseline but made aware afterwards. Researchers conducting clinical interviews to assess relapse risk, and MRI scans, were masked to treatment allocation during the whole trial.

#### - Instructions for experimental conditions

In resting-state, participants were told to close their eyes and relax. During rumination, participants were guided through a rumination induction adapted for eyes closed. The paradigm has been validated (Karl et al., 2018) and is known to induce negative self-related thoughts in individuals with a history of recurrent depression. Instructions consisted in first rehearsing a sad autobiographical memory, and subsequently staying in that sad mood, reflecting on self-related causes and consequences of the low mood. Participants were free to choose their own memory, and the choice was not restricted over the sessions.

#### - Magnetic resonance imaging acquisition

Brain imaging was acquired using a 3T Siemens Magnetom Skyra 3T scanner (Siemens Healthineers/Erlangen, Germany, software version Scout) with a 32-channel head coil. A structural three-dimensional T1-weighted (3D-T1) scan was acquired with the following parameters: 176 slices covering the whole brain; echo time (TE)= 3.8 ms; repetition time (TR)= 2300 ms; inversion time=31260 ms; flip angle=8°; field of view (FOV)= 256 mm; spatial resolution=1x1x1 mm³; Generalized Autocalibrating Partially Parallel Acquisitions (GRAPPA)= 2, phase-encoding direction= AP. Functional data was collected in all scan conditions (resting-state, mindfulness, resting-state and rumination induction) obtaining for each 203 volumes of 2D gradient-echo EPI fMRI data with the following parameters: 52 ascending axial slices covering the whole brain; 3.8 x 3.8 x 3.8 mm³; FOV=192; GRAPPA= 2; Multiband= 2; TE=30 ms; TR=1480 ms; flip angle=65°; phase-encoding direction=AP.

### - BOLD fMRI pre-processing

Preprocessing was done using FSL tools (version 6.0) (Smith et al., 2004) with standard procedures. The following were included: skull-stripping (BET tool (Smith, 2002)) with functional to structural image registering (FLIRT tool (Jenkinson et al., 2002) with default settings for Boundary-Based registration); structural image to standard space registering (FNIRT tool with default settings for 12 degrees of freedom and warp-resolution of 10 mm); motion correction (MCFLIRT tool (Jenkinson et al., 2002)); spatial smoothing using a 5mm kernel. Motion correction was done using an independent component analysis-based strategy for Automatic Removal of Motion Artifacts (ICA-AROMA (Pruim et al., 2015)). For further denoising, the first five eigenvariates of time courses extracted from white matter and cerebrospinal fluid masks (segmentation done with FAST tool (Zhang et al., 2001)) were removed (using *fsl_glm*). As a last step, data was high-pass filtered (100 seconds cut-off).

### - Brain parcellation

Neuroimaging data was parcellated into 90 brain areas using the Automated Anatomical Labelling (van Aalderen et al.) atlas (Tzourio-Mazoyer et al., 2002).

For each cortical brain area, we have the number of voxels overlapping the 7 well-known Yeo networks (Visual, Somatomotor, Dorsal Attention, Salience, Limbic, Control and Default Mode Networks) (Yeo et al., 2011). When applicable, we associate a brain area to the resting-state network in which it contains more voxels. All subcortical brain areas are linked directly to the SCN.

### - Whole-brain computational model

#### *Hopf bifurcation model*

We used a Stuart-Landau oscillator, corresponding to the normal form of a supercritical Hopf bifurcation, to represent the local dynamics of each brain area. The mathematical expression of the whole-brain dynamics coupled via a connectivity matrix *C* is the following (Deco, 2024):

$$\frac{dz_j}{dt} = (a_j + i\omega_j)z_j - |z_j|^2 z_j + \sum_{k=1}^{N} C_{jk}(z_k - z_j) + \eta_j. \quad (1)$$

Here, $z_j$ is the complex variable denoting the state of a node $j$ ($z_j = x_j + iy_j$), and $\eta_j$ is Gaussian noise with variance $\sigma^2$. Furthermore, $\omega_j$ is the node frequency, calculated by applying a band-pass filter of 0.008-0.08 Hz to the fMRI signals.

Lastly, the bifurcation parameter $a_j$ governs the dynamics, in which if $a_j > 0$ the local dynamics generate self-sustained oscillations of frequency $f_j = w_j/2\pi$, settling into a stable limit cycle. If $a_j <$

0, the local dynamics represent a stable spiral point where noisy or damped oscillations are generated if noise is present or not, respectively. The fMRI signals are modeled by the real part of the state variables (i.e., $x_j = Real(z_j)$).

***Hopf linear approximation***

It has been shown that when the bifurcation parameter is slightly negative ($a_j = -0.02$) it preserves dynamically responding brain networks and resting-state network structure, and fluctuating stochastically structured signals are generated (Sanz Perl et al., 2023). This allows linearizing the dynamics and finding an analytical solution for the functional connectivity matrix $C$ (i.e., Pearson correlations between brain areas) (Ponce-Alvarez & Deco, 2024). By implementing a linear noise approximation **Equation 1** is expressed in vector form as:

$$\frac{dz}{dt} = (a - S + i\omega) \odot z - (z \odot \bar{z})z + Cz + \eta. \tag{2}$$

Here, $z = [z_1, \ldots, z_N]^T$, $a = [a_1, \ldots, a_N]^T$, $\omega = [\omega_1, \ldots, \omega_N]^T$ and $\eta = [\eta_1, \ldots, \eta_N]^T$. Furthermore, $S = [S_1, \ldots, S_N]^T$ indicates the connectivity strength of each node (i.e., $S_i = \sum_j C_{ij}$). Lastly, $[\,]^T$ is the transpose operation, $\odot$ corresponds to the element-wise product, and $\bar{z}$ is the complex conjugate of $z$.

The equation describes the linear fluctuations around the fixed point $z = 0$, the solution of $\frac{dz}{dt} = 0$. By separating the real and imaginary parts of the state variables and discarding higher-order terms (i.e., $(z \odot \bar{z})z$), the evolution of the linear fluctuations follow a Langevin stochastic linear equation:

$$\frac{d}{dt}\delta u = J\delta u + \eta. \tag{3}$$

In this equation, $\delta u = [\delta x, \delta y]^{\mathrm{T}} = [\delta x_1, \ldots, \delta x_N, \delta y_1, \ldots, \delta y_N]^{\mathrm{T}}$ is a 2N-dimensional vector containing the fluctuations of the real and imaginary state variables. In addition, $J$ is a $2N\,X\,2N$ Jacobian matrix of the system evaluated at the fixed point $z = 0$. It has the following form as block matrix:

$$J = \begin{bmatrix} Jxx & Jxy \\ Jyx & Jyy \end{bmatrix}. \tag{4}$$

Here, the matrices $J_{xx}$, $J_{xy}$, $J_{yx}$ and $J_{yy}$ have a size of $N\,X\,N$; $J_{xx} = J_{yy} = diag(a - S) + C$ and $J_{xy} = -J_{yx} = diag(\omega)$, in which $diag(v)$ corresponds to the diagonal matrix with the vector $v$ as the diagonal. The linearization is possible only if $z = 0$ is a stable solution of the system, which corresponds to $J$ having all eigenvalues with a negative real part.

The covariance matrix $K = \langle \delta u \delta u^T \rangle$ is calculated by expressing **Equation 3** as $d\delta u = J\delta u dt + dW$. Here, $dW$ is a 2N-dimensional Wiener process, whose covariance is $\langle dW dW^T \rangle = Qdt$, and $Q$ is the noise covariance matrix (which is diagonal if the noise is uncorrelated). Using Itô's stochastic calculus, $d(\delta u \delta u^T) = d(\delta u)\delta u^T + \delta u d(\delta u^T) + d(\delta u)d(\delta u^T)$. And, taking expectations, noting $\langle \delta u dW^T \rangle = 0$, and keeping terms to first order in the differential $dt$, the following equation is obtained:

$$\frac{dK}{dt} = JK + KJ^T + Q. \tag{5}$$

Solving this equation for the case $\frac{dK}{dt} = 0$, the stationary covariance is obtained analytically using the eigen-decomposition of the Jacobian matrix $J$ (Deco & Kringelbach, 2014).

***Model optimisation – Generative Effective Connectivity***

We optimised the coupling connectivity matrix $C$ until best fit between the model and empirical data using asymmetrical measures. We used as a starting point a standard structural connectivity matrix, the diffusion MRI data of the public repository of the Human Connectome Project (Van Essen et al., 2013). Then, we applied a pseudo-gradient procedure until convergence to a final matrix which is the generative effective connectivity (GEC).

We updated each known existing pair of anatomical connections including homotopic areas (i.e., brain areas in opposite hemispheres) given tractography captures them less accurately. This was obtained by adding the difference between the simulated and empirical functional correlation matrix and normalised time-shifted covariance matrix as follows:

$$C_{ij} = C_{ij} + \alpha\left(FC_{ij}^{emp} - FC_{ij}^{sim}\right) + \varsigma\left(FS_{ij}^{emp}(\tau) - FS_{ij}^{sim}(\tau)\right), \tag{6}$$

where $\alpha$ equals 0.0004 and $\varsigma$ is set to 0.0001. The empirical functional connectivity ($FC^{emp}$) is computed with the normalised covariance matrix of the timeseries. The empirical normalised time-shifted covariance ($FS^{emp}(\tau)$) is obtained by dividing each pair of regions *(i,j)* from the shifted covariance matrix ($KS^{emp}(\tau)$) by $\sqrt{KS_{ii}^{emp}(0)KS_{jj}^{emp}(0)}$. With respect to the simulated matrices, the simulated functional connectivity ($FC^{sim}$) is obtained from the first $N$ rows and columns of the simulated covariance matrix $K$, corresponding to the real part of the dynamics and representing the BOLD fMRI signal. The simulated normalised time-shifted covariance ($FS^{sim}(\tau)$) is computed by selecting the first $N$ rows and columns of the simulated time-shifted covariance ($KS^{sim}(\tau)$), and dividing each pair of regions $(i,j)$ by $\sqrt{KS_{ii}^{sim}(0)KS_{jj}^{sim}(0)}$. Here,

$$KS^{sim}(\tau) = exp(\tau J)K, \tag{7}$$

where $KS^{sim}(0) = K$.

We used these time-shifted matrices (time-shift set to $\tau = 2$), to break the symmetry during the optimisation process and obtain a GEC which captures the asymmetry intrinsic to hierarchical organisation and enhance fitting quality (Kringelbach et al., 2023). We initialised the procedure to a total of 1500 iterations, and every 100 iterations we computed the difference between the empirical and modelled measures. If it was higher than the difference from the previous iteration, or smaller than 0.001 from the last iteration, the optimisation procedure finished. We first calculated the GEC at a group level for each treatment (MBCT+TAU and TAU), condition (resting-state and rumination) and session (before and after) by using the DTI template as a starting point. Then, we calculated the GEC at an individual level, using the GEC of the participant's group as a starting point. We averaged the peak frequency of each brain area across all individuals of a specific group.

**- Inside-out framework**

We implemented the INSIDEOUT framework developed by (Deco et al., 2022) to the empirical data. This way, we could evaluate the arrow of time by comparing the causal relationship between the forward pairwise time series and their reversed backward version.

Let's consider a timeseries $x(t)$ evolving from an initial state $A_1$ to a final state $A_2$, and another timeseries $y(t)$ evolving from $B_1$ to $B_2$. Their reversed backward versions, $x^{(r)}(t)$ and $y^{(r)}(t)$, can be obtained by flipping their time ordering (from $A_2/B_2$ to $A_1/B_1$). Then, the causal dependency between $x(t)$ and $y(t)$ can be measured with their time-shifted correlation. In the forward evolution, this is given by

$$c_{forward}(\Delta t) = < x(t), y(t + \Delta t) >, \tag{8}$$

and in the reversed backward evolution, by

$$c_{reversal}(\Delta t) = < x^{(r)}(t), y^{(r)}(t + \Delta t). \tag{9}$$

Then, the absolute difference of the forward and reversed causal dependencies between the timeseries, at a time shift Δt=T, can be calculated as

$$I_{x,t}(T) = \left|c_{forward}(T) - c_{reversal}(T)\right|. \tag{10}$$

For the multidimensional case, the forward and reversal matrices of time-shifted correlations can be defined as follows. Let's denote the forward version as $x_i(t)$, reflecting the dynamical evolution of the variable describing the system, and $x_i^{(r)}(t)$ the corresponding reversed backward version. The sub-index $i$ denotes the dimensions of the system.

Then, the forward and reversal matrices are given, respectively, by

$$FS_{forward,ij}(\Delta t) = -\frac{1}{2} log\left(1 - < x_i(t), x_j(t + \Delta t) >^2\right), \tag{11}$$

$$FS_{reversal,ij}(\Delta t) = -\frac{1}{2} log\left(1 - < x_i{}^{(r)}(t), x_j{}^{(r)}(t + \Delta t) >^2\right). \tag{12}$$

Lastly, their quadratic distance can be calculated at a given shift Δt=T as

$$I = ||FS_{forward}(\mathrm{T}) - FS_{reversal}(\mathrm{T})||_2. \quad (13)$$

The notation $||Q||_2$ is defined as the mean of the absolute squares of the elements of the matrix $Q$. Thus, a matrix $FS_{diff}$, whose elements are the squared difference of each pair of elements of the forward and reversal matrices, can be defined as

$$FS_{diff}ij = (FS_{forward}ij(\mathrm{T}) - FS_{reversal}ij(\mathrm{T}))^2. \quad (14)$$

Therefore, I is the mean value of the elements of $FS_{diff}$.

We used an optimal T=2 following the model-based analysis.

**Participants clinical and behavioural scores**

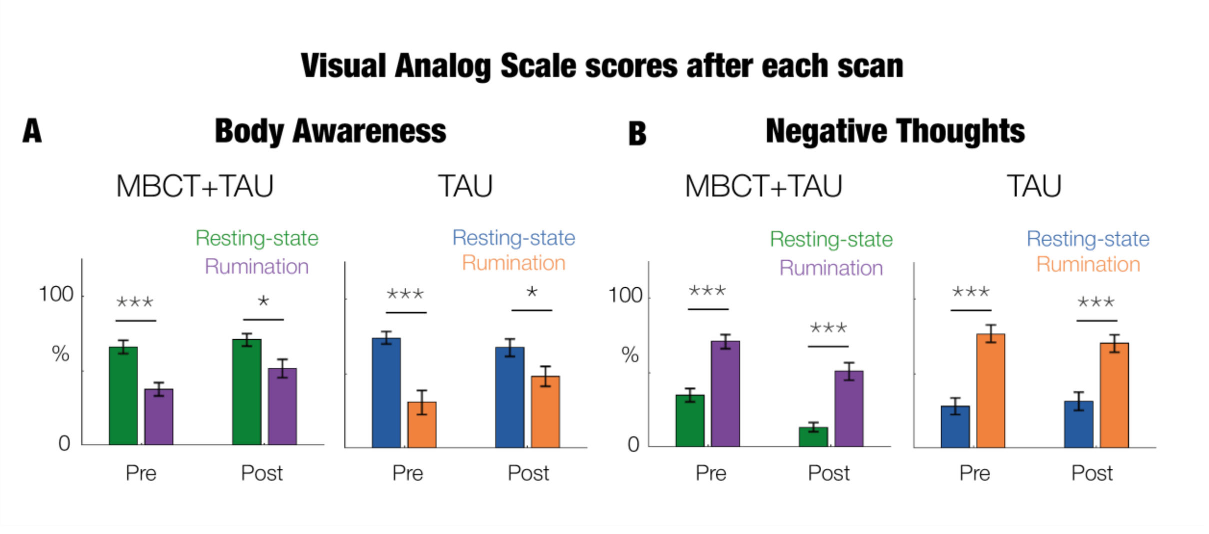


***Supplementary Figure S1. Visual Analog Scale scores after each scan comparing resting-state and rumination.*** *Participants had to rate on a computer screen in the scanner from 0-100% in a Visual Analog Scale (VAS) their body awareness ("I was aware of my body") and negative thoughts ("I had negative thoughts about myself"). Differences between conditions (RS, resting-state, and RUM, rumination) for each treatment (MBCT+TAU and TAU) and session (before and after). Barplots show the mean across participants and errorbars show their standard error of the mean. Significance is represented by asterisks (***, $p < 0.001$; *, $p < 0.05$). There was one participant with no response for the rumination TAU group.*

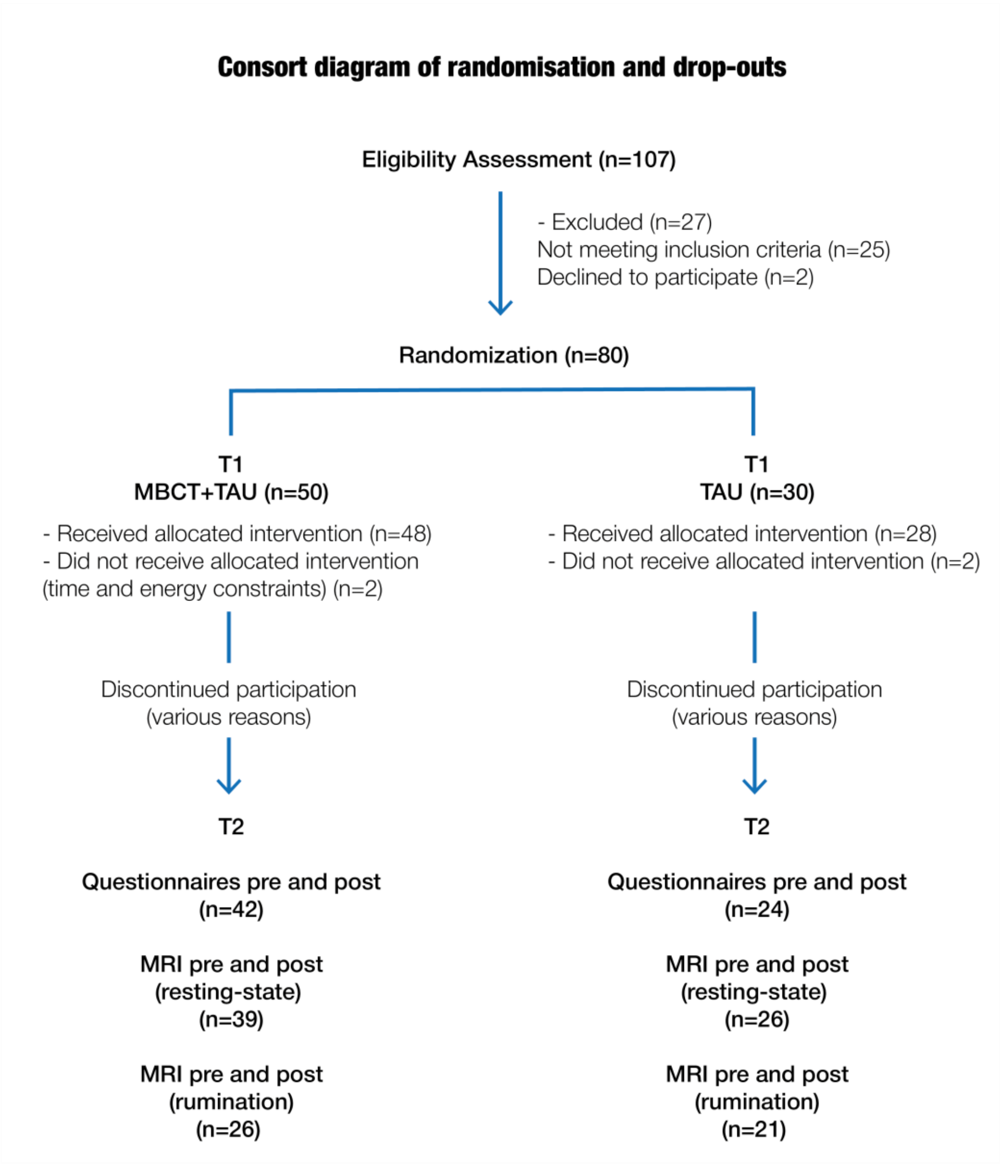


***Supplementary Figure S2. Consort diagram of randomisation and drop-outs adapted from (van der Velden et al., 2023).***

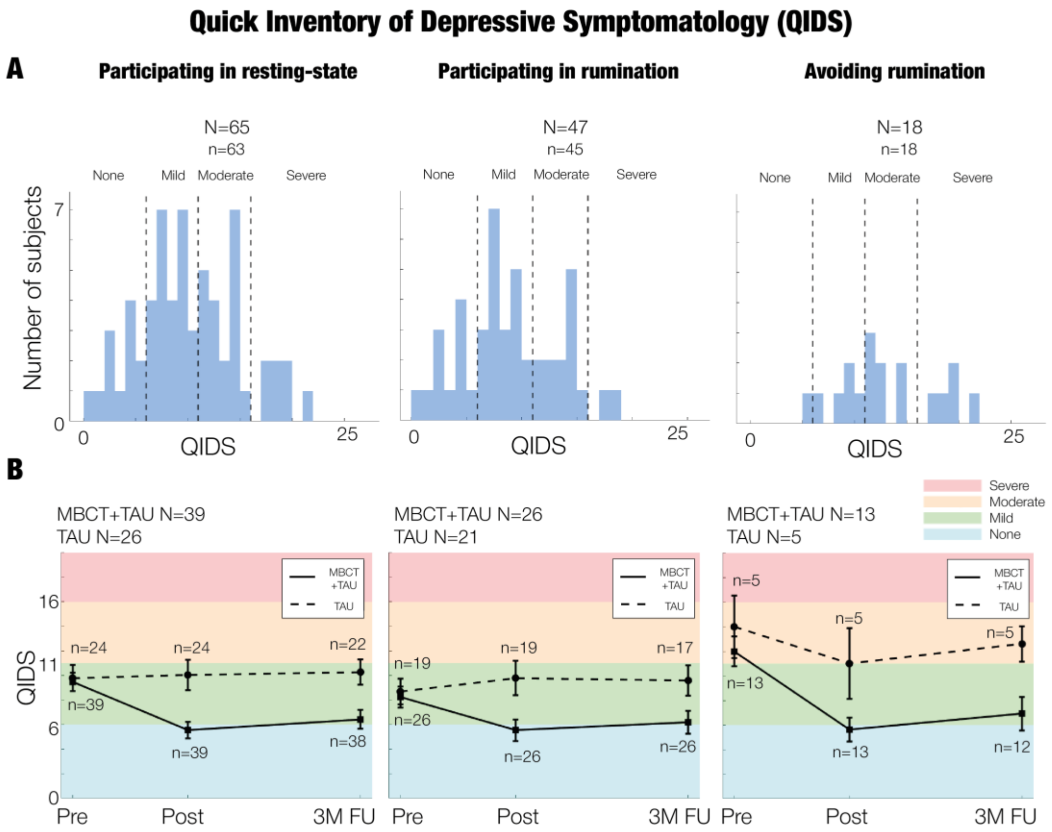


***Supplementary Figure S3. Depressive symptoms measured by Quick Inventory of Depressive Symptoms (QIDS) across time and group.*** *The columns correspond to the group of participants that performed resting-state, rumination, and those that avoided the rumination task. We show the total number of participants with available fMRI data (N) and the ones with available data for this clinical analysis (n).* ***A.*** *Histograms show the depressive symptoms at baseline.* ***B.*** *Lineplots show the QIDS scores at baseline, post-, and 3 months follow-up (FU) (x-axis). The lines are the mean, and the error bars the standard error of the mean for both MBCT+TAU and TAU (dashed line). Colour-coding represents asymptomatic for QIDS<6 (blue), mild symptoms for QIDS 6-10 (green), moderate symptoms for QIDS 11-15 (orange) and severe symptoms for QIDS for QIDS 16-25 (red).*

| Category | Participating in resting-state | | | | | | | | | | Participating in rumination | | | | | | | | | |
|---|---|---|---|---|---|---|---|---|---|---|---|---|---|---|---|---|---|---|---|---|
| | MBCT + TAU | | | | | TAU | | | | | MBCT + TAU | | | | | TAU | | | | |
| | value | min | max | std | N | value | min | max | std | N | value | min | max | std | N | value | min | max | std | N |
| **Sociodemographic characteristics** | | | | | | | | | | | | | | | | | | | | |
| **Age** | 44.513 | 18 | 68 | 13.076 | 39 | 44.808 | 24 | 70 | 12.103 | 26 | 41.115 | 18 | 66 | 13.483 | 26 | 42.952 | 24 | 66 | 11.647 | 21 |
| **Sex (M/F)** | 9/30 | - | - | - | 39 | 4/22 | - | - | - | 26 | 6/20 | - | - | - | 26 | 4/17 | - | - | - | 21 |
| **Clinical characteristics** | | | | | | | | | | | | | | | | | | | | |
| **Symptomatic** | 84.615 | - | - | - | 39 | 75.000 | - | - | - | 24 | 80.769 | - | - | - | 26 | 68.421 | - | - | - | 19 |
| **mADM** | 87.179 | - | - | - | 39 | 80.769 | - | - | - | 26 | 80.769 | - | - | - | 26 | 80.952 | - | - | - | 21 |
| **Childhood trauma** | 37.344 | 25 | 83 | 13.879 | 32 | 42.038 | 25 | 97 | 16.880 | 26 | 39.333 | 25 | 83 | 15.285 | 21 | 39.524 | 26 | 90 | 13.310 | 21 |
| **Treatment outcomes** | | | | | | | | | | | | | | | | | | | | |
| **QIDS** | 9.487 | 1 | 19 | 4.662 | 39 | 9.792 | 0 | 21 | 5.167 | 24 | 8.231 | 1 | 18 | 4.375 | 26 | 8.684 | 0 | 17 | 4.547 | 19 |
| **FFMQ** | 43.054 | 27 | 63 | 7.863 | 37 | 45.077 | 33 | 61 | 8.202 | 26 | 42.840 | 27 | 56 | 7.403 | 25 | 45.905 | 33 | 61 | 8.130 | 21 |
| **PSS** | 21.405 | 11 | 34 | 6.099 | 37 | 20.731 | 9 | 32 | 6.792 | 26 | 20.760 | 11 | 31 | 6.287 | 25 | 19.571 | 9 | 31 | 6.439 | 21 |
| **RRS** | 53.543 | 29 | 73 | 10.489 | 37 | 57.115 | 44 | 71 | 8.368 | 26 | 55.120 | 29 | 73 | 10.879 | 25 | 56.714 | 44 | 71 | 8.361 | 21 |
| **EQ** | 31.130 | 21 | 48 | 6.701 | 37 | 31.500 | 19 | 45 | 7.208 | 26 | 30.400 | 21 | 43 | 5.752 | 25 | 32.362 | 19 | 45 | 7.121 | 21 |
| **MAIA NO** | 12.865 | 4 | 18 | 2.800 | 37 | 14.154 | 7 | 20 | 3.295 | 26 | 12.440 | 4 | 18 | 3.110 | 25 | 13.762 | 7 | 20 | 3.434 | 21 |
| **MAIA ND** | 9.351 | 4 | 15 | 2.720 | 37 | 8.962 | 3 | 14 | 2.474 | 26 | 9.800 | 5 | 15 | 2.754 | 25 | 9.048 | 3 | 14 | 2.598 | 21 |
| **MAIA EA** | 15.225 | 7.5 | 23.75 | 3.596 | 37 | 16.635 | 11.25 | 25 | 4.370 | 26 | 15.133 | 7.5 | 23.75 | 4.073 | 25 | 16.250 | 11.25 | 25 | 4.108 | 21 |
| **MAIA AR** | 17.243 | 8 | 29 | 5.085 | 37 | 17.769 | 9 | 32 | 5.172 | 26 | 16.440 | 8 | 29 | 5.463 | 25 | 17.571 | 9 | 32 | 5.427 | 21 |
| **MAIA BL** | 6.649 | 3 | 11 | 2.003 | 37 | 7.346 | 3 | 15 | 3.358 | 26 | 6.840 | 3 | 11 | 2.192 | 25 | 7.143 | 3 | 15 | 3.425 | 21 |
| **MAIA TR** | 9.027 | 3 | 15 | 3.078 | 37 | 8.462 | 3 | 15 | 3.591 | 26 | 8.880 | 3 | 15 | 3.321 | 25 | 8.429 | 3 | 15 | 3.842 | 21 |

***Supplementary Table S1. Baseline characteristics per treatment group and engagement in the rumination task.*** *Abbreviations: ADM, maintenance antidepressant medication; QIDS, Quick Inventory of Depressive Symptomatology; FFMQ, Five Factor Mindfulness Questionnaire; PSS, Perceived Stress Scale; RRS, Rumination Response Scale; EQ, Experience Questionnaire; MAIA, Multidimensional Assessment of Interoceptive Awareness; NO, noticing; ND, not-distracting; EA, emotional awareness; AR, attention regulation; BL, body listening; TR, trusting.*

**Main Analysis**

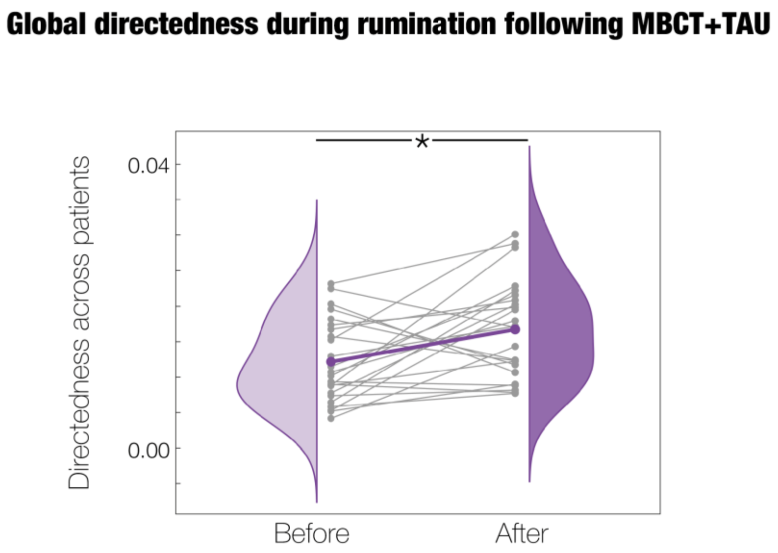


***Supplementary Figure S4. Global hierarchical reconfiguration during rumination with MBCT+TAU.*** *At a global level, significant differences in directedness between the before and after treatment (*, p < 0.05) when removing two subjects with consistent extreme positive values across sessions. Grey lines represent the trajectory of each individual whereas the coloured line represents their averaged trajectory.*

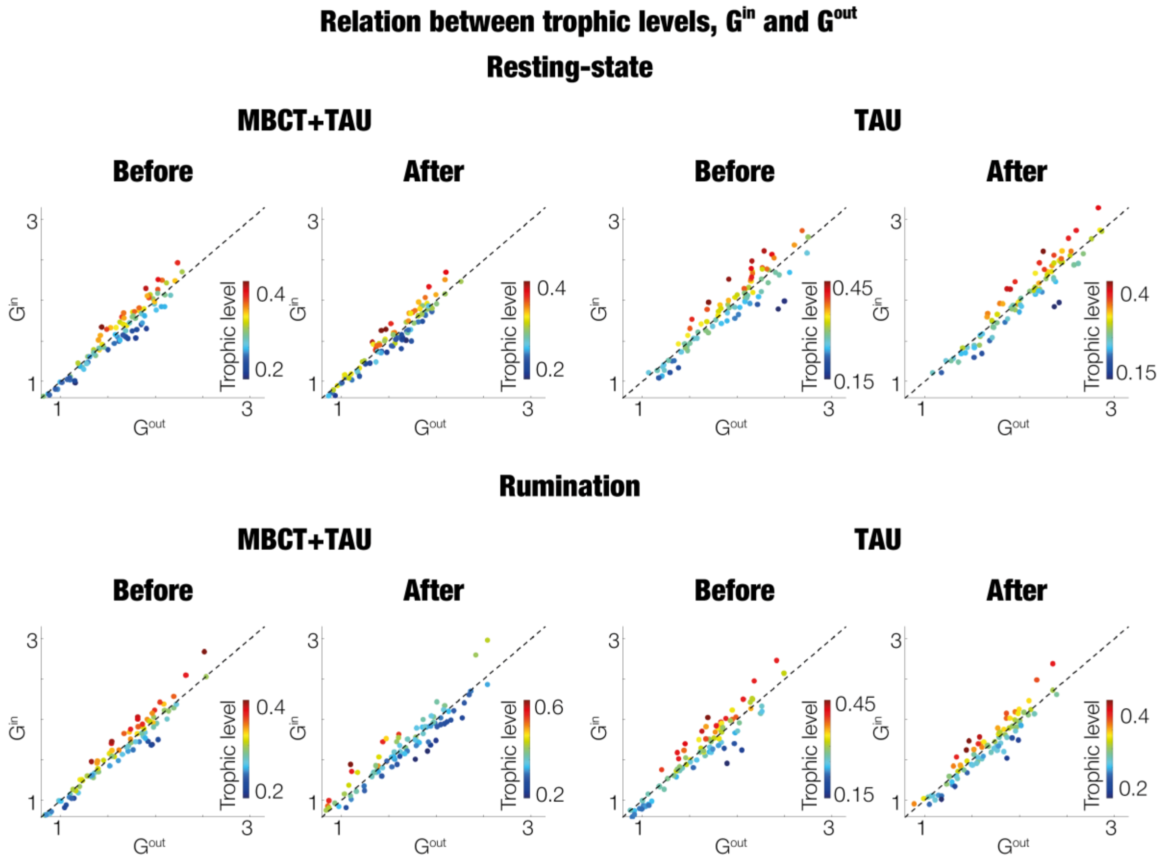


***Supplementary Figure S5. Relation between brain area trophic level and in- and out- GEC.*** *There is one plot for each scan condition (resting-state, rumination), treatment (MBCT+TAU, TAU) and session (before, after). Each graph shows brain areas scattered by the sum of the in-flow (y-axis) and sum of out-flow (x-axis) of the GEC, all with the same axis range. Brain areas are colour-coded by their trophic level, adjusting resolution in each plot to clearly identify the top (in red) and bottom areas (in blue). The dotted diagonal is included to identify data-points (i.e., areas) with in>out (above diagonal) and in<out (below diagonal).*

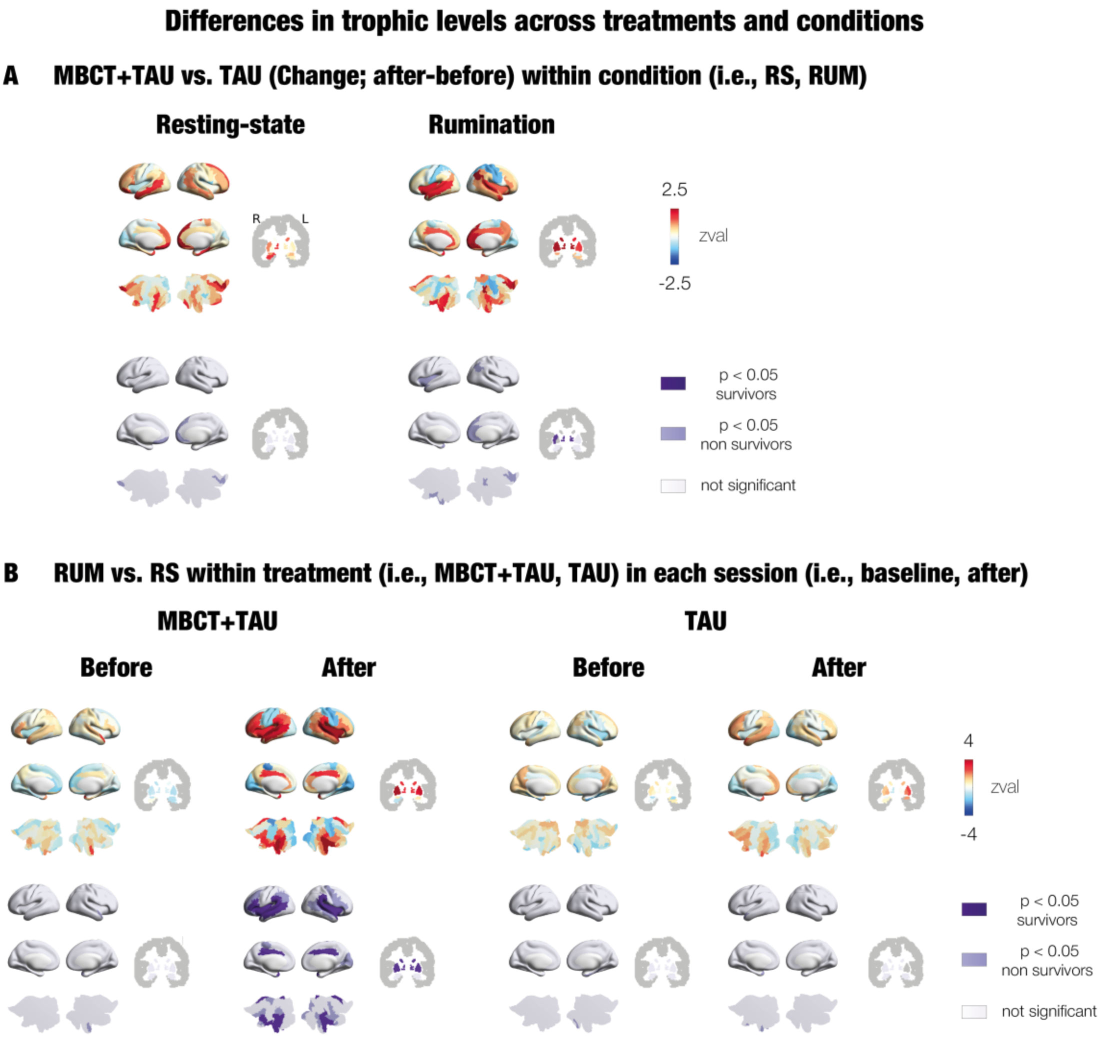


***Supplementary Figure S6. Statistics between hierarchical levels of treatments and conditions.*** *We computed statistics between two groups of individuals for each brain area. In each panel, the first row of brain renders show the z-value of the statistics. The second row of brain renders show significance, with dark purple for significant brain areas (p < 0.05) surviving correction by multiple comparisons, lilac for significant brain areas (p < 0.05) not surviving correction by multiple comparisons, and white for brain areas which are non-significant. Subcortical regions are shown on slices in Montreal Neurological Institute (MNI) space (coronal axis y= -6 mm).* ***A.*** *Treatment comparison (MBCT+TAU vs. TAU) for the change (after-before) within each scan condition (resting-state left, rumination right). Positive z-values mean more positive changes (or less negative changes) in MBCT+TAU compared to TAU. The opposite occurs for negative z-values, revealing stronger negative changes (or less positive changes) in MBCT+TAU compared to TAU. In both resting-state and rumination, approximately 5% of areas were significant, none surviving correction by multiple comparisons.* ***B.*** *Condition comparison (rumination vs. resting-state) for each treatment (MBCT+TAU and TAU) and session (before and after). Positive z-values mean higher hierarchical levels for rumination compared to resting-state. For MBCT+TAU before, TAU before and TAU after, approximately 5% of areas were significant, none surviving correction by multiple comparisons. For MBCT+TAU after treatment, approximately 40% of brain areas were significant, 25% surviving correction for multiple comparisons.*

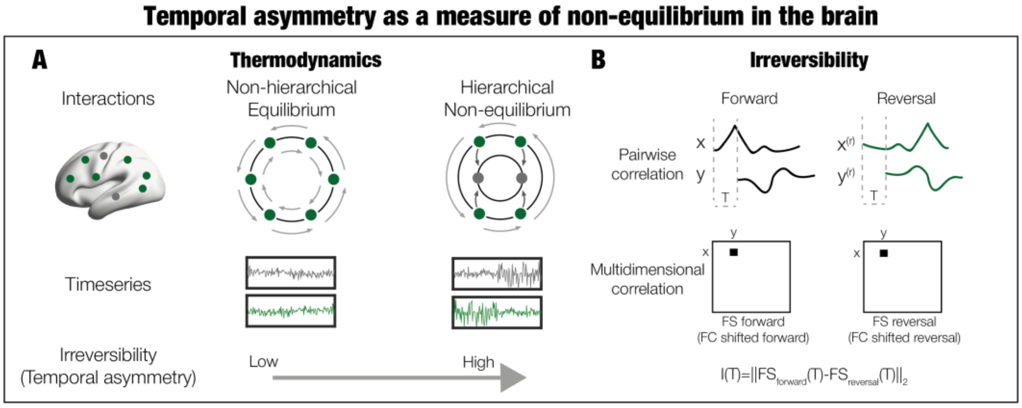


***Supplementary Figure S7. The hierarchical organisation of the brain can be measured indirectly by the empirical irreversibility of the timeseries. A.*** *Two systems of circles and arrows illustrating states and transitions, respectively. A flat hierarchy is characterised by low irreversibility and a high hierarchy is less reversible over time.* ***B.*** *Irreversibility can be computed in a multidimensional case such as the brain by calculating the distance between the forward and backwards time-shifted correlation matrices at a given shift in time.*

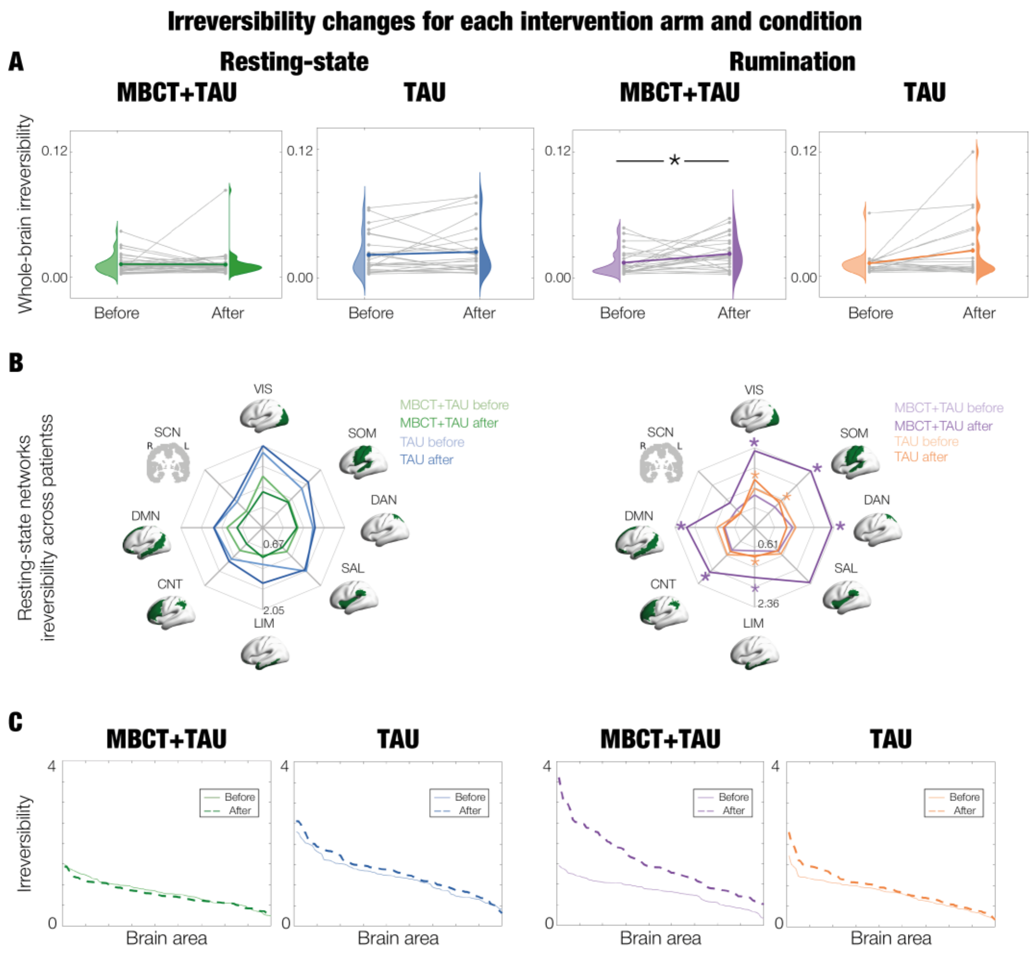


***Supplementary Figure S8. Irreversibility changes with treatment.*** *Throughout the whole figure results are shown for resting-state in the left column and for rumination in the right column.* ***A.*** *At a global level, significant differences between before and after treatment were found only in MBCT+TAU during rumination (*, p < 0.05). Gray lines represent the trajectory of each individual whereas the coloured line represents their averaged trajectory within each intervention arm and condition. We also computed statistics at baseline, revealing no significant differences between treatments and scan conditions except for MBCT+TAU resting-state vs. TAU resting-state (p < 0.05).* ***B.*** *Spider plots show the weighted irreversibility by resting-state network. For each network, firstly, we multiplied the number of voxels of each area belonging to the network by the area's irreversibility value. Then, we added the resulting values across all areas and divided by the total sum of voxels for that network. Significant changes are represented by asterisks (*, p < 0.05), in bold surviving correction by multiple comparisons. For rumination MBCT+TAU, all networks changed significantly except for the SAL and SCN, and from the significant networks all survived correction for multiple comparisons except the LIM. For rumination TAU, significant changes were found in the VIS, SOM and LIM, non surviving correction for multiple comparisons.* ***C.*** *The median irreversibility at a regional level across patients is ordered in descending order. The brain area ordering (x-axis) is different for each scan condition and treatment. After MBCT+TAU treatment, in rumination, the irreversibility increased, and the steepness of the slope also increased.*

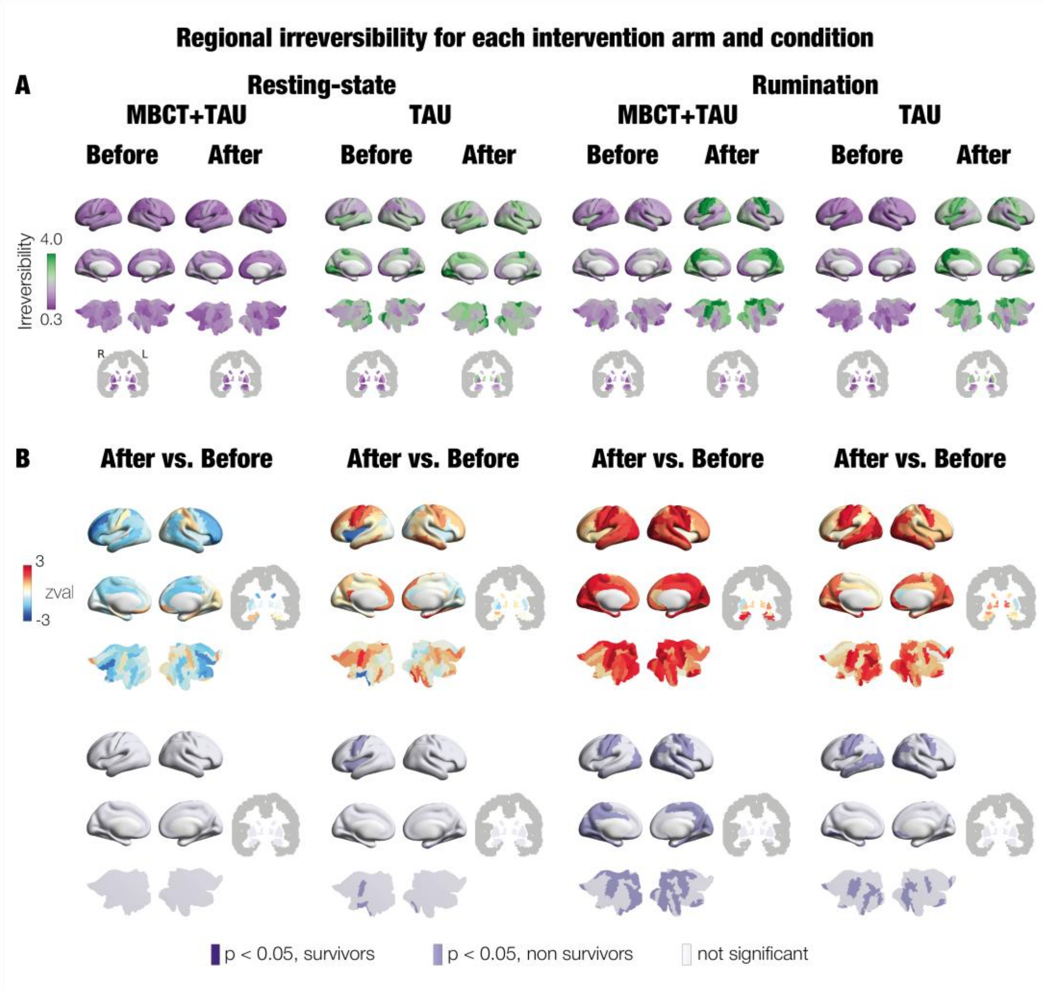


***Supplementary Figure S9. Regional irreversibility changes with treatment. A.*** *Brain renders show the average irreversibility across individuals for each scan condition (resting-state and rumination), treatment (MBCT+TAU and TAU) and session (before and after).* ***B.*** *Differences in irreversibility levels between before and after treatment of each condition and treatment. The first row shows the z-values with the colourscale representing positive values in red and negative in blue. The second row shows the significance, with p < 0.05 in purple, in dark the ones surviving correction by multiple comparisons and light (i.e., lilac) the ones not surviving. Subcortical regions are shown on slices in Montreal Neurological Institute (MNI) space (coronal axis y= -6 mm).*

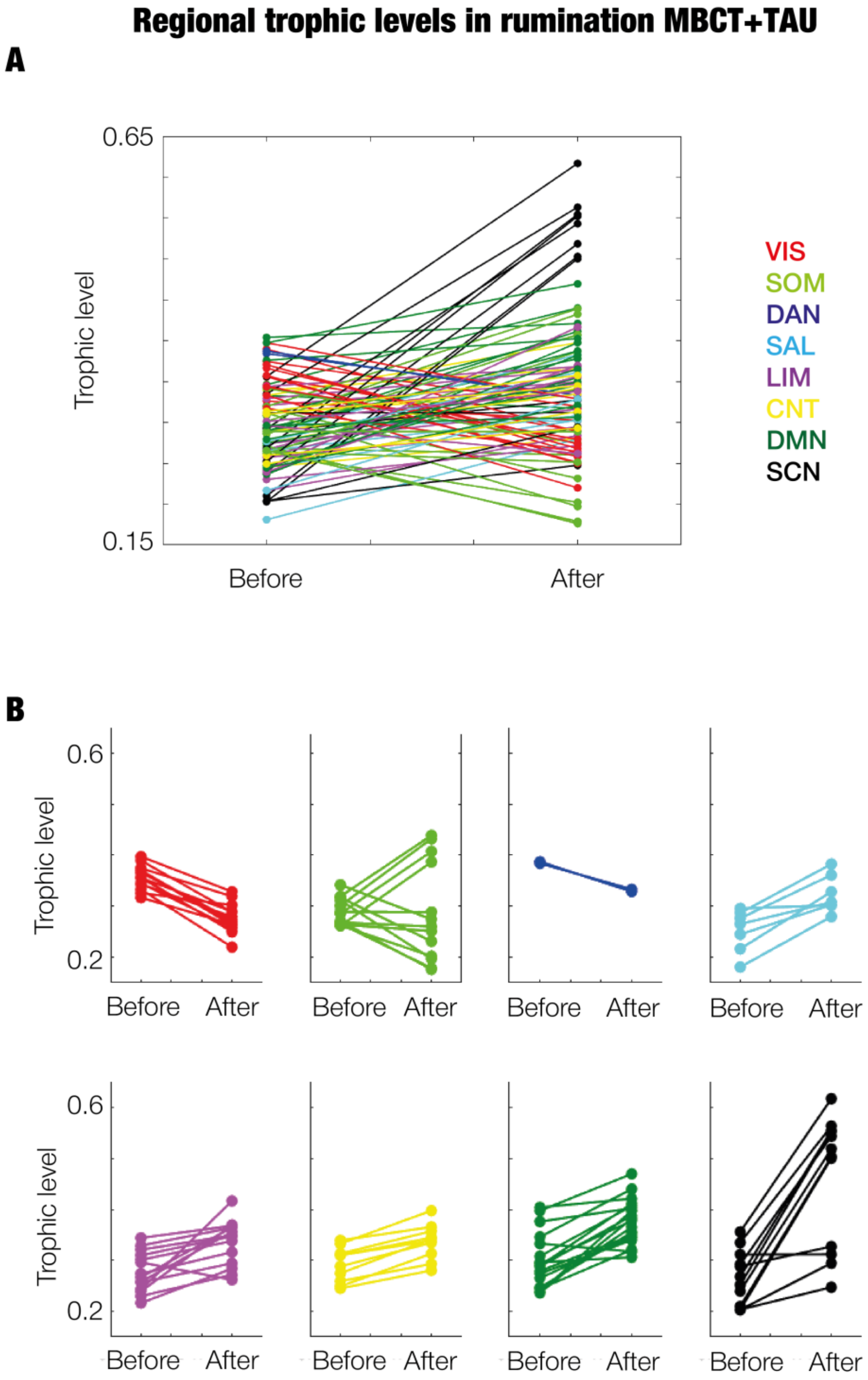


***Supplementary Figure S10. Changes of regional trophic levels across patients in MBCT+TAU rumination associated with Yeo resting-state networks.*** *Plots showing the trophic level of each brain area averaged across patients for before and after MBCT+TAU in rumination. The colour-coding represents the Yeo resting-state network (RSN) (Yeo et al., 2011) a brain area has highest contribution to. The purpose is for visual inspection, and no statistics are performed* ***A.*** *Plot shows all brain areas together.* ***B.*** *Each plot shows brain areas corresponding to the RSN with highest contribution.*

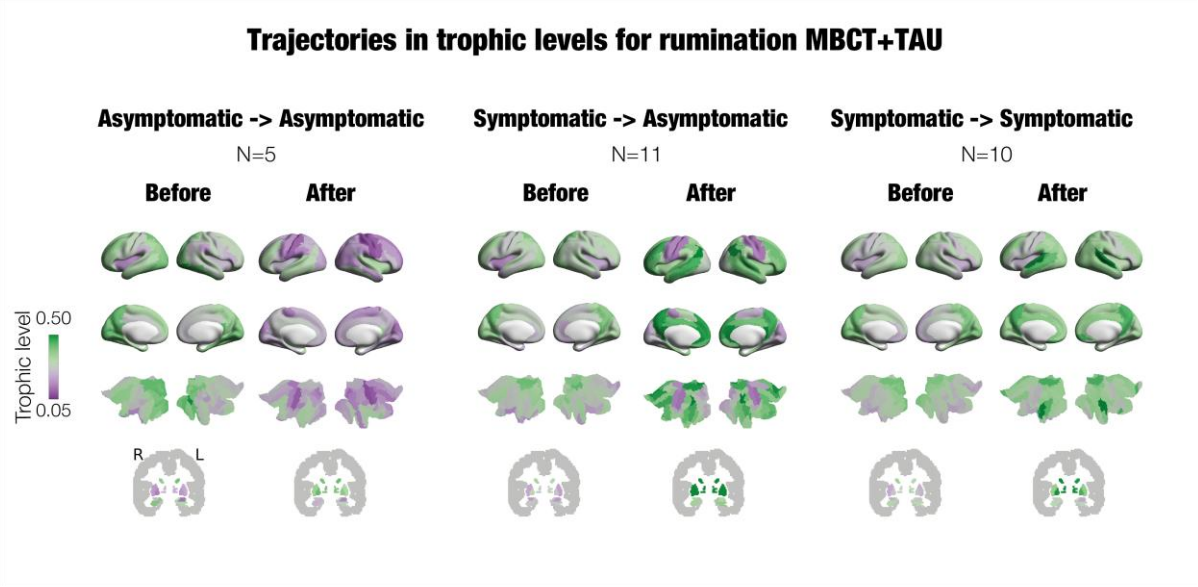


***Supplementary Figure S11. Regional trophic levels in rumination MBCT+TAU for different trajectories in depressive symptoms.*** *The trajectories correspond to* ***A.*** *from asymptomatic to asymptomatic (N=5 individuals),* ***B.*** *from symptomatic to asymptomatic (N=11 individuals with N=2 moderate to none, and N=9 mild to none) and* ***C.*** *symptomatic to symptomatic (N=10 individuals with N=1 severe to mild, N=3 moderate to mild, N=4 mild to mild, and N=2 mild to severe). For each trajectory in each session (before and after), the average trophic levels across patients are rendered on brain maps. Green shows higher trophic levels whereas purple corresponds to lower trophic levels. The scaling of colorbar is adjusted to facilitate recognition of changes within this analysis. Subcortical regions are shown on slices in Montreal Neurological Institute (MNI) space (coronal axis y= -6 mm).*

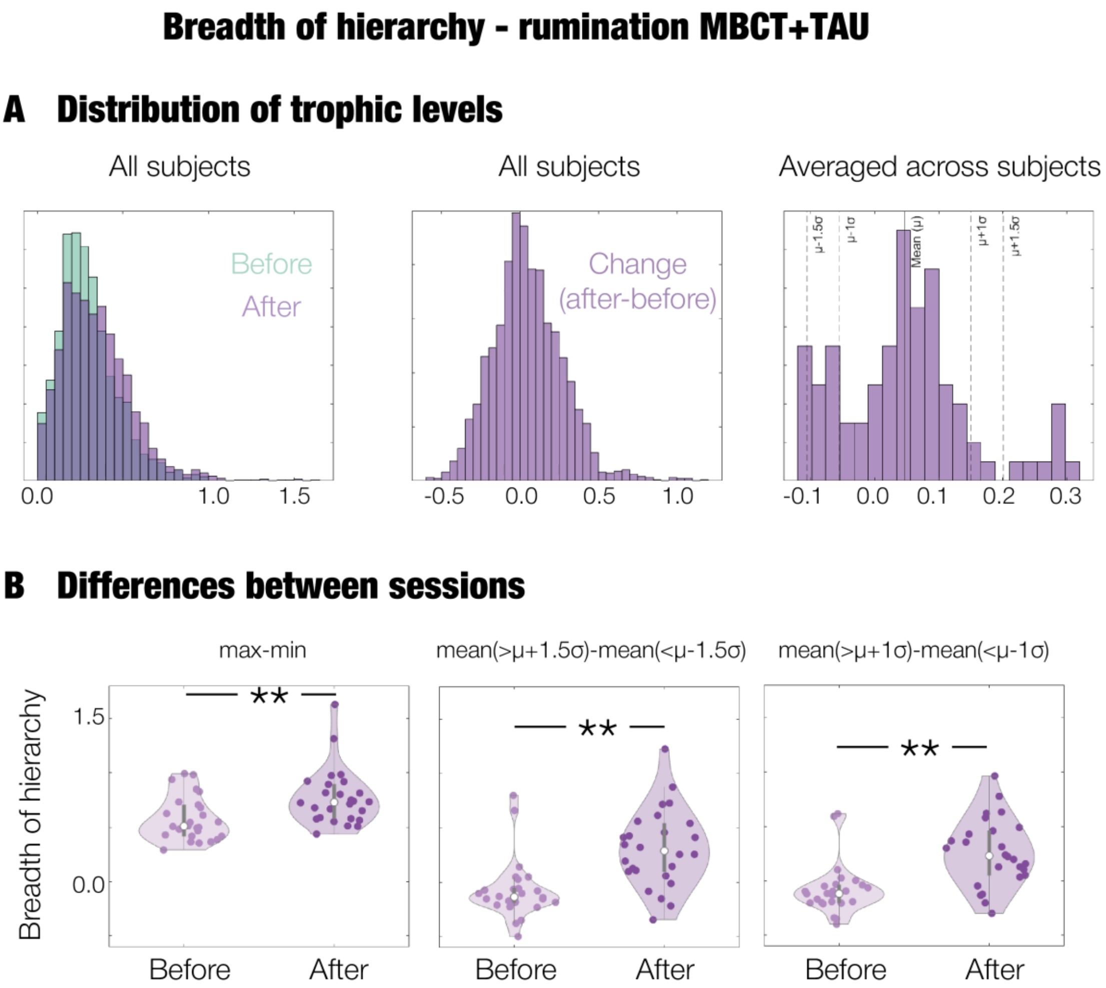


***Supplementary Figure S12.*** *Breadth of hierarchy for MBCT+TAU during rumination.* ***A.*** *Histograms showing the distribution of trophic levels: before and after treatment for all subjects (left); change (after-before) for all subjects (middle); change (after-before) averaged across subjects (right). This last plot includes the thresholds for the top and bottom 1 and 1.5 standard deviations from the mean.* ***B.*** *Each plot shows the difference between before and after treatment for the breadth of hierarchy calculated as: maximum minus the minimum trophic level (left); first identifying the areas above and below a threshold of 1.5 standard deviations of the mean (13% of regions), computing their averages separately, and calculating the top mean minus the bottom mean (middle); first identifying the areas above and below a threshold of 1 standard deviations of the mean (32% of regions), computing their averages separately, and calculating the top mean minus the bottom mean (right). Significance is represented by asterisks (**, p < 0.01). The right plot shows highest significance, obtained when using as threshold 1 standard deviation, which also included a larger proportion of regions. The specific regions at the top end for this case are: left and right thalamus, pallidum, putamen, caudate; left anterior cingulate and right olfactory cortex. The bottom areas are: left and right calcarine, cuneus, lingual; left and right superior, middle and inferior occipital cortex; left and right precentral, postcentral, paracentral lobules; right superior parietal cortex.*

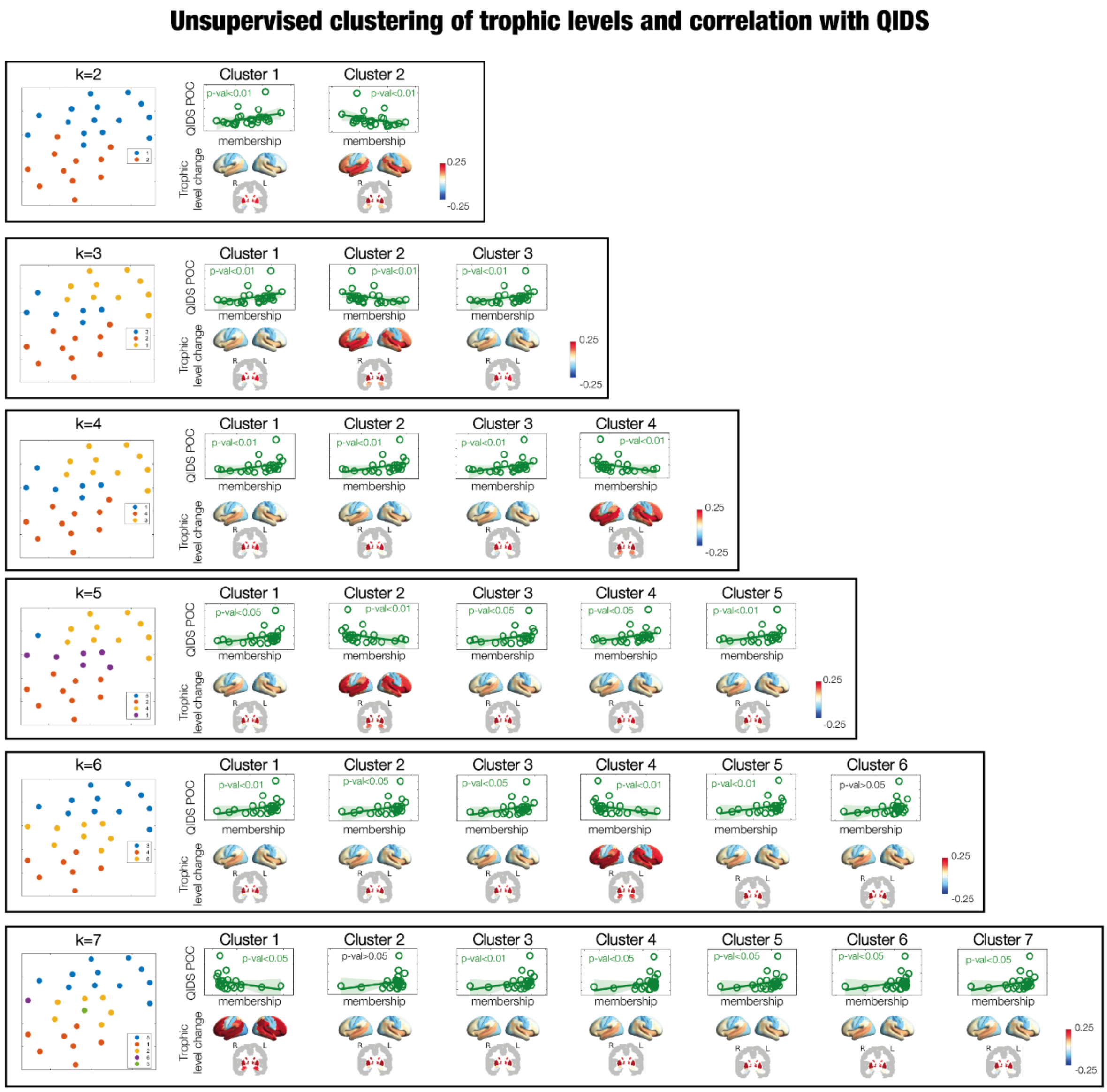


***Supplementary Figure S13. Fingerprint of change (after-before) in trophic levels associated with depressive score improvement in MBCT+TAU rumination.*** *Following the methodological pipeline from (Dagnino et al., 2023), we implemented an unsupervised clustering to the trophic level change to evaluate if we could uncover a brain prototype of response. We clustered the change in trophic levels for the rumination MBCT+TAU group using fuzzy c-means algorithm and cluster centres k ranging from 2-7. Each data-point corresponds to an individual with 90 features (i.e., 90 trophic levels of all brain areas). Fuzzy c-means is a clustering algorithm which assigns data a membership value to each cluster centre, shown to work well for convex datasets and show robustness to outliers. It has the benefit of being non-binary, and unsupervised. Each row shows the results of the clustering in a given number of cluster centres. The plots in the left show the high-dimensional data visualised in a 2D plot with t-distributed stochastic neighbour embedding (t-SNE) and coloured in the cluster centre with highest membership value. Then, for each cluster centroid in a given clustering configuration (i.e., row), we calculated the correlation between the membership to each cluster, with the percentage of change (POC) in clinical score QIDS calculated as (after-before)/before. Furthermore, we constructed brain renders with the features (i.e., trophic levels) of each cluster centroid. Subcortical regions are shown*

*on slices in Montreal Neurological Institute (MNI) space (coronal axis y= -6 mm). The brain renders of all cluster centres display similar regional reconfigurations, with all brain areas revealing positive changes except to those highly linked to the VIS and SOM. In all clustering results across k partitions (ranging from 2 to 7), there is one specific centroid with a significantly negative correlation with the POC in behavioural score. In other words, the higher the membership of a patient to this given centroid, the more negative the QIDS POC (i.e., improved in clinical severity of depression). The rendering of trophic levels for this cluster shows highest strength for the areas changing positively.*

| | RS MBCT+TAU BEFORE | | | RS MBCT+TAU AFTER | | |
|---|---|---|---|---|---|---|
| Top 20% | 54. Occipital_Inf_R | VIS | 0.421 | 72. Caudate_R | SCN | 0.407 |
| | 43. Calcarine_L | VIS | 0.406 | 36. Cingulum_Post_R | DMN | 0.407 |
| | 48. Lingual_R | VIS | 0.394 | 71. Caudate_L | SCN | 0.397 |
| | 67. Precuneus_L | DMN | 0.382 | 67. Precuneus_L | DMN | 0.382 |
| | 53. Occipital_Inf_L | VIS | 0.379 | 68. Precuneus_R | DMN | 0.378 |
| | 36. Cingulum_Post_R | DMN | 0.375 | 54. Occipital_Inf_R | VIS | 0.370 |
| | 45. Cuneus_L | VIS | 0.371 | 35. Cingulum_Post_L | DMN | 0.365 |
| | 49. Occipital_Sup_L | VIS | 0.371 | 23. Frontal_Sup_Medial_L | DMN | 0.362 |
| | 47. Lingual_L | VIS | 0.366 | 25. Frontal_Med_Orb_L | DMN | 0.354 |
| | 50. Occipital_Sup_R | VIS | 0.365 | 26. Frontal_Med_Orb_R | DMN | 0.353 |
| | 68. Precuneus_R | DMN | 0.363 | 38. Hippocampus_R | SCN | 0.353 |
| | 44. Calcarine_R | VIS | 0.363 | 77. Thalamus_L | SCN | 0.351 |
| | 72. Caudate_R | SCN | 0.361 | 43. Calcarine_L | VIS | 0.351 |
| | 60. Parietal_Sup_R | DAN | 0.348 | 24. Frontal_Sup_Medial_R | DMN | 0.348 |
| | 70. Paracentral_Lobule_R | SOM | 0.346 | 78. Thalamus_R | SCN | 0.343 |
| | 59. Parietal_Sup_L | DAN | 0.346 | 48. Lingual_R | VIS | 0.342 |
| | 90. Temporal_Inf_R | LIM | 0.345 | 53. Occipital_Inf_L | VIS | 0.341 |
| | 69. Paracentral_Lobule_L | SOM | 0.341 | 3. Frontal_Sup_L | DMN | 0.337 |
| Bottom 20% | 27. Rectus_L | LIM | 0.225 | 79. Heschl_L | SOM | 0.221 |
| | 42. Amygdala_R | SCN | 0.224 | 14. Frontal_Inf_Tri_R | CNT | 0.221 |
| | 79. Heschl_L | SOM | 0.222 | 80. Heschl_R | SOM | 0.218 |
| | 28. Rectus_R | LIM | 0.221 | 12. Frontal_Inf_Oper_R | CNT | 0.215 |
| | 80. Heschl_R | SOM | 0.220 | 81. Temporal_Sup_L | SOM | 0.215 |
| | 13. Frontal_Inf_Tri_L | CNT | 0.219 | 88. Temporal_Pole_Mid_R | LIM | 0.215 |
| | 12. Frontal_Inf_Oper_R | CNT | 0.215 | 73. Putamen_L | SCN | 0.214 |
| | 15. Frontal_Inf_Orb_L | DMN | 0.215 | 84. Temporal_Pole_Sup_R | LIM | 0.207 |
| | 84. Temporal_Pole_Sup_R | LIM | 0.210 | 57. Postcentral_L | SOM | 0.207 |
| | 30. Insula_R | SAL | 0.208 | 74. Putamen_R | SCN | 0.205 |
| | 11. Frontal_Inf_Oper_L | CNT | 0.207 | 18. Rolandic_Oper_R | SOM | 0.203 |
| | 14. Frontal_Inf_Tri_R | CNT | 0.205 | 63. SupraMarginal_L | SAL | 0.198 |
| | 83. Temporal_Pole_Sup_L | LIM | 0.200 | 17. Rolandic_Oper_L | SOM | 0.196 |
| | 88. Temporal_Pole_Mid_R | LIM | 0.196 | 64. SupraMarginal_R | SAL | 0.191 |
| | 29, Insula_L | SAL | 0.190 | 11. Frontal_Inf_Oper_L | CNT | 0.185 |
| | 73. Putamen_L | SCN | 0.188 | 30. Insula_R | SAL | 0.185 |
| | 74. Putamen_R | SCN | 0.186 | 41. Amygdala_L | SCN | 0.183 |
| | 87. Temporal_Pole_Mid_L | LIM | 0.167 | 29, Insula_L | SAL | 0.170 |

***Supplementary Table S2. Trophic levels of each session for MBCT+TAU during resting-state.*** *The table shows the brain areas with highest 20% (first row) and lowest 20% trophic levels before (left column) and after (right column) treatment. Each brain area is assigned the resting-state network in which it shows the greatest contribution, and its trophic level.*

| | RS TAU BEFORE | | | RS TAU AFTER | | |
|---|---|---|---|---|---|---|
| Top 20% | 54. Occipital_Inf_R | VIS | 0.454 | 43. Calcarine_L | VIS | 0.424 |
| | 53. Occipital_Inf_L | VIS | 0.449 | 36. Cingulum_Post_R | DMN | 0.403 |
| | 43. Calcarine_L | VIS | 0.440 | 54. Occipital_Inf_R | VIS | 0.401 |
| | 48. Lingual_R | VIS | 0.440 | 67. Precuneus_L | DMN | 0.392 |
| | 47. Lingual_L | VIS | 0.423 | 48. Lingual_R | VIS | 0.386 |
| | 49. Occipital_Sup_L | VIS | 0.421 | 68. Precuneus_R | DMN | 0.382 |
| | 44. Calcarine_R | VIS | 0.413 | 72. Caudate_R | SCN | 0.376 |
| | 70. Paracentral_Lobule_R | SOM | 0.409 | 45. Cuneus_L | VIS | 0.375 |
| | 45. Cuneus_L | VIS | 0.397 | 53. Occipital_Inf_L | VIS | 0.375 |
| | 69. Paracentral_Lobule_L | SOM | 0.393 | 44. Calcarine_R | VIS | 0.373 |
| | 60. Parietal_Sup_R | DAN | 0.390 | 47. Lingual_L | VIS | 0.370 |
| | 50. Occipital_Sup_R | VIS | 0.390 | 46. Cuneus_R | VIS | 0.357 |
| | 67. Precuneus_L | DMN | 0.389 | 35. Cingulum_Post_L | DMN | 0.353 |
| | 56. Fusiform_R | VIS | 0.387 | 49. Occipital_Sup_L | VIS | 0.349 |
| | 46. Cuneus_R | VIS | 0.385 | 69. Paracentral_Lobule_L | SOM | 0.348 |
| | 90. Temporal_Inf_R | LIM | 0.381 | 70. Paracentral_Lobule_R | SOM | 0.347 |
| | 55. Fusiform_L | VIS | 0.379 | 56. Fusiform_R | VIS | 0.339 |
| | 68. Precuneus_R | DMN | 0.376 | 60. Parietal_Sup_R | DAN | 0.339 |
| Bottom 20% | 16. Frontal_Inf_Orb_R | DMN | 0.250 | 64. SupraMarginal_R | SAL | 0.226 |
| | 31. Cingulum_Ant_L | DMN | 0.248 | 63. SupraMarginal_L | SAL | 0.224 |
| | 83. Temporal_Pole_Sup_L | LIM | 0.247 | 14. Frontal_Inf_Tri_R | CNT | 0.223 |
| | 78. Thalamus_R | SCN | 0.247 | 28. Rectus_R | LIM | 0.221 |
| | 15. Frontal_Inf_Orb_L | DMN | 0.245 | 30. Insula_R | SAL | 0.221 |
| | 13. Frontal_Inf_Tri_L | CNT | 0.243 | 41. Amygdala_L | SCN | 0.220 |
| | 75. Pallidum_L | SCN | 0.242 | 21. Olfactory_L | LIM | 0.220 |
| | 30. Insula_R | SAL | 0.236 | 15. Frontal_Inf_Orb_L | DMN | 0.219 |
| | 14. Frontal_Inf_Tri_R | CNT | 0.229 | 29, Insula_L | SAL | 0.217 |
| | 32. Cingulum_Ant_R | DMN | 0.228 | 13. Frontal_Inf_Tri_L | CNT | 0.213 |
| | 79. Heschl_L | SOM | 0.223 | 83. Temporal_Pole_Sup_L | LIM | 0.205 |
| | 21. Olfactory_L | LIM | 0.220 | 11. Frontal_Inf_Oper_L | CNT | 0.198 |
| | 76. Pallidum_R | SCN | 0.209 | 27. Rectus_L | LIM | 0.191 |
| | 29, Insula_L | SAL | 0.209 | 76. Pallidum_R | SCN | 0.187 |
| | 87. Temporal_Pole_Mid_L | LIM | 0.200 | 87. Temporal_Pole_Mid_L | LIM | 0.181 |
| | 88. Temporal_Pole_Mid_R | LIM | 0.189 | 88. Temporal_Pole_Mid_R | LIM | 0.174 |
| | 74. Putamen_R | SCN | 0.159 | 73. Putamen_L | SCN | 0.126 |
| | 73. Putamen_L | SCN | 0.150 | 74. Putamen_R | SCN | 0.123 |

***Supplementary Table S3. Trophic levels of each session for TAU during resting-state.*** *The table shows the brain areas with highest 20% (first row) and lowest 20% trophic levels before (left column) and after (right column) treatment. Each brain area is assigned the resting-state network in which it shows the greatest contribution, and its trophic level.*

| | RUM MBCT+TAU BEFORE | | | RUM MBCT+TAU AFTER | | |
|---|---|---|---|---|---|---|
| Top 20% | 67. Precuneus_L | DMN | 0.404 | 72. Caudate_R | SCN | 0.617 |
| | 36. Cingulum_Post_R | DMN | 0.397 | 71. Caudate_L | SCN | 0.563 |
| | 54. Occipital_Inf_R | VIS | 0.397 | 78. Thalamus_R | SCN | 0.554 |
| | 43. Calcarine_L | VIS | 0.390 | 76. Pallidum_R | SCN | 0.552 |
| | 60. Parietal_Sup_R | DAN | 0.387 | 77. Thalamus_L | SCN | 0.543 |
| | 59. Parietal_Sup_L | DAN | 0.384 | 75. Pallidum_L | SCN | 0.518 |
| | 68. Precuneus_R | DMN | 0.376 | 73. Putamen_L | SCN | 0.503 |
| | 53. Occipital_Inf_L | VIS | 0.374 | 74. Putamen_R | SCN | 0.500 |
| | 49. Occipital_Sup_L | VIS | 0.371 | 36. Cingulum_Post_R | DMN | 0.470 |
| | 50. Occipital_Sup_R | VIS | 0.370 | 35. Cingulum_Post_L | DMN | 0.440 |
| | 48. Lingual_R | VIS | 0.366 | 82. Temporal_Sup_R | SOM | 0.439 |
| | 44. Calcarine_R | VIS | 0.357 | 81. Temporal_Sup_L | SOM | 0.432 |
| | 72. Caudate_R | SCN | 0.356 | 67. Precuneus_L | DMN | 0.421 |
| | 45. Cuneus_L | VIS | 0.356 | 22. Olfactory_R | LIM | 0.417 |
| | 51. Occipital_Mid_L | VIS | 0.356 | 85. Temporal_Mid_L | DMN | 0.411 |
| | 35. Cingulum_Post_L | DMN | 0.346 | 80. Heschl_R | SOM | 0.407 |
| | 47. Lingual_L | VIS | 0.345 | 65. Angular_L | DMN | 0.407 |
| | 90. Temporal_Inf_R | LIM | 0.344 | 68. Precuneus_R | DMN | 0.402 |
| Bottom 20% | 25. Frontal_Med_Orb_L | DMN | 0.247 | 48. Lingual_R | VIS | 0.274 |
| | 11. Frontal_Inf_Oper_L | CNT | 0.245 | 87. Temporal_Pole_Mid_L | LIM | 0.272 |
| | 64. SupraMarginal_R | SAL | 0.245 | 50. Occipital_Sup_R | VIS | 0.268 |
| | 16. Frontal_Inf_Orb_R | DMN | 0.244 | 56. Fusiform_R | VIS | 0.264 |
| | 26. Frontal_Med_Orb_R | DMN | 0.244 | 49. Occipital_Sup_L | VIS | 0.262 |
| | 22. Olfactory_R | LIM | 0.243 | 40. ParaHippocampal_R | LIM | 0.262 |
| | 75. Pallidum_L | SCN | 0.240 | 17. Rolandic_Oper_L | SOM | 0.260 |
| | 28. Rectus_R | LIM | 0.239 | 47. Lingual_L | VIS | 0.258 |
| | 31. Cingulum_Ant_L | DMN | 0.238 | 20. Supp_Motor_Area_R | SOM | 0.251 |
| | 32. Cingulum_Ant_R | DMN | 0.236 | 45. Cuneus_L | VIS | 0.251 |
| | 87. Temporal_Pole_Mid_L | LIM | 0.230 | 44. Calcarine_R | VIS | 0.250 |
| | 30. Insula_R | SAL | 0.217 | 41. Amygdala_L | SCN | 0.247 |
| | 88. Temporal_Pole_Mid_R | LIM | 0.217 | 69. Paracentral_Lobule_L | SOM | 0.231 |
| | 73. Putamen_L | SCN | 0.210 | 46. Cuneus_R | VIS | 0.220 |
| | 41. Amygdala_L | SCN | 0.204 | 1. Precentral_L | SOM | 0.202 |
| | 42. Amygdala_R | SCN | 0.203 | 58. Postcentral_R | SOM | 0.197 |
| | 74. Putamen_R | SCN | 0.203 | 2. Precentral_R | SOM | 0.179 |
| | 29, Insula_L | SAL | 0.181 | 57. Postcentral_L | SOM | 0.176 |

***Supplementary Table S4. Trophic levels of each session for MBCT+TAU during rumination.*** *The table shows the brain areas with highest 20% (first row) and lowest 20% trophic levels before (left column) and after (right column) treatment. Each brain area is assigned the resting-state network in which it shows the greatest contribution, and its trophic level.*

| | RUM TAU BEFORE | | | RUM TAU AFTER | | |
|---|---|---|---|---|---|---|
| Top 20% | 43. Calcarine_L | VIS | 0.455 | 71. Caudate_L | SCN | 0.430 |
| | 53. Occipital_Inf_L | VIS | 0.432 | 36. Cingulum_Post_R | DMN | 0.429 |
| | 36. Cingulum_Post_R | DMN | 0.424 | 72. Caudate_R | SCN | 0.415 |
| | 60. Parietal_Sup_R | DAN | 0.416 | 67. Precuneus_L | DMN | 0.398 |
| | 44. Calcarine_R | VIS | 0.414 | 60. Parietal_Sup_R | DAN | 0.392 |
| | 54. Occipital_Inf_R | VIS | 0.414 | 35. Cingulum_Post_L | DMN | 0.389 |
| | 45. Cuneus_L | VIS | 0.411 | 68. Precuneus_R | DMN | 0.378 |
| | 68. Precuneus_R | DMN | 0.411 | 43. Calcarine_L | VIS | 0.378 |
| | 48. Lingual_R | VIS | 0.411 | 23. Frontal_Sup_Medial_L | DMN | 0.372 |
| | 67. Precuneus_L | DMN | 0.410 | 3. Frontal_Sup_L | DMN | 0.365 |
| | 47. Lingual_L | VIS | 0.406 | 70. Paracentral_Lobule_R | SOM | 0.364 |
| | 70. Paracentral_Lobule_R | SOM | 0.398 | 9. Frontal_Mid_Orb_L | CNT | 0.363 |
| | 50. Occipital_Sup_R | VIS | 0.395 | 75. Pallidum_L | SCN | 0.362 |
| | 49. Occipital_Sup_L | VIS | 0.386 | 7. Frontal_Mid_L | CNT | 0.359 |
| | 46. Cuneus_R | VIS | 0.384 | 6. Frontal_Sup_Orb_R | LIM | 0.357 |
| | 69. Paracentral_Lobule_L | SOM | 0.381 | 10. Frontal_Mid_Orb_R | CNT | 0.354 |
| | 59. Parietal_Sup_L | DAN | 0.371 | 19. Supp_Motor_Area_L | SAL | 0.353 |
| | 55. Fusiform_L | VIS | 0.368 | 59. Parietal_Sup_L | DAN | 0.352 |
| Bottom 20% | 63. SupraMarginal_L | SAL | 0.229 | 73. Putamen_L | SCN | 0.260 |
| | 39. ParaHippocampal_L | LIM | 0.229 | 18. Rolandic_Oper_R | SOM | 0.260 |
| | 15. Frontal_Inf_Orb_L | DMN | 0.227 | 79. Heschl_L | SOM | 0.260 |
| | 14. Frontal_Inf_Tri_R | CNT | 0.221 | 28. Rectus_R | LIM | 0.251 |
| | 87. Temporal_Pole_Mid_L | LIM | 0.221 | 17. Rolandic_Oper_L | SOM | 0.248 |
| | 12. Frontal_Inf_Oper_R | CNT | 0.218 | 78. Thalamus_R | SCN | 0.246 |
| | 79. Heschl_L | SOM | 0.215 | 12. Frontal_Inf_Oper_R | CNT | 0.242 |
| | 42. Amygdala_R | SCN | 0.213 | 14. Frontal_Inf_Tri_R | CNT | 0.242 |
| | 41. Amygdala_L | SCN | 0.212 | 21. Olfactory_L | LIM | 0.241 |
| | 40. ParaHippocampal_R | LIM | 0.210 | 39. ParaHippocampal_L | LIM | 0.238 |
| | 88. Temporal_Pole_Mid_R | LIM | 0.210 | 87. Temporal_Pole_Mid_L | LIM | 0.231 |
| | 75. Pallidum_L | SCN | 0.209 | 42. Amygdala_R | SCN | 0.229 |
| | 76. Pallidum_R | SCN | 0.207 | 64. SupraMarginal_R | SAL | 0.226 |
| | 64. SupraMarginal_R | SAL | 0.205 | 30. Insula_R | SAL | 0.211 |
| | 29, Insula_L | SAL | 0.190 | 40. ParaHippocampal_R | LIM | 0.192 |
| | 30. Insula_R | SAL | 0.180 | 29, Insula_L | SAL | 0.190 |
| | 74. Putamen_R | SCN | 0.155 | 74. Putamen_R | SCN | 0.184 |
| | 73. Putamen_L | SCN | 0.129 | 88. Temporal_Pole_Mid_R | LIM | 0.168 |

***Supplementary Table S5. Trophic levels of each session for TAU during rumination.*** *The table shows the brain areas with highest 20% (first row) and lowest 20% trophic levels before (left column) and after (right column) treatment. Each brain area is assigned the resting-state network in which it shows the greatest contribution, and its trophic level.*

| Brain area | RSN | Z-value | P-value | Correction |
|---|---|---|---|---|
| 26. Frontal_Med_Orb_R | DMN | 3.028234548 | 0.002459871 | x |
| 28. Rectus_R | LIM | 2.986369554 | 0.002823113 | x |
| 78. Thalamus_R | SCN | 2.958459559 | 0.003091808 | x |
| 23. Frontal_Sup_Medial_L | DMN | 2.916594565 | 0.003538754 | x |
| 25. Frontal_Med_Orb_L | DMN | 2.763089588 | 0.005725705 | x |
| 24. Frontal_Sup_Medial_R | DMN | 2.735179592 | 0.006234625 | x |
| 49. Occipital_Sup_L | VIS | -2.721224594 | 0.006504056 | x |
| 27. Rectus_L | LIM | 2.661167587 | 0.007787019 | x |
| 45. Cuneus_L | VIS | -2.567719617 | 0.010236992 | x |
| 32. Cingulum_Ant_R | DMN | 2.483989629 | 0.012991959 | x |
| 59. Parietal_Sup_L | DAN | -2.288619659 | 0.022101461 | x |
| 85. Temporal_Mid_L | DMN | 2.204889671 | 0.027461837 | x |
| 71. Caudate_L | SCN | 2.190934673 | 0.02845652 | x |
| 50. Occipital_Sup_R | VIS | -2.190934673 | 0.02845652 | x |
| 1. Precentral_L | SOM | -2.093249688 | 0.036326871 | x |
| 31. Cingulum_Ant_L | DMN | 2.051384694 | 0.040229498 | x |
| 60. Parietal_Sup_R | DAN | -2.051384694 | 0.040229498 | x |
| 2. Precentral_R | SOM | -1.995564702 | 0.045981325 | x |
| 57. Postcentral_L | SOM | -1.981609704 | 0.04752294 | x |

***Supplementary Table S6. Trophic level change in MBCT+TAU resting-state.*** *Brain areas significant change between after vs. before, with the assigned resting-state network in which it shows the greatest contribution, z-value, p-value and if they survive correction by multiple comparisons (✓) or not (x).*

| Brain area | RSN | Z-value | P-value | Correction |
|---|---|---|---|---|
| 53. Occipital_Inf_L | VIS | -2.22232301 | 0.026261484 | x |
| 58. Postcentral_R | SOM | -2.22232301 | 0.026261484 | x |
| 41. Amygdala_L | SCN | -2.120731101 | 0.033944438 | x |
| 40. ParaHippocampal_R | LIM | -2.095333124 | 0.03614139 | x |
| 27. Rectus_L | LIM | -2.044537169 | 0.040900509 | x |
| 89. Temporal_Inf_L | LIM | -2.044537169 | 0.040900509 | x |
| 11. Frontal_Inf_Oper_L | CNT | -1.993741215 | 0.046180343 | x |
| 90. Temporal_Inf_R | LIM | -1.968343238 | 0.049028558 | x |
| 10. Frontal_Mid_Orb_R | CNT | -1.968343238 | 0.049028558 | x |

***Supplementary Table S7. Trophic level change in TAU resting-state.*** *Brain areas significant change between after vs. before, with the assigned resting-state network in which it shows the greatest contribution, z-value, p-value and if they survive correction by multiple comparisons (✓) or not (x).*

| Brain area | RSN | Z-value | P-value | Correction |
|---|---|---|---|---|
| 78. Thalamus_R | SCN | 4.177967259 | 2.94126E-05 | ✓ |
| 77. Thalamus_L | SCN | 4.000181418 | 6.32939E-05 | ✓ |
| 74. Putamen_R | SCN | 3.847793555 | 0.000119186 | ✓ |
| 76. Pallidum_R | SCN | 3.7969976 | 0.000146459 | ✓ |
| 75. Pallidum_L | SCN | 3.695405691 | 0.000219536 | ✓ |
| 73. Putamen_L | SCN | 3.568415805 | 0.000359146 | ✓ |
| 72. Caudate_R | SCN | 3.543017828 | 0.000395576 | ✓ |
| 71. Caudate_L | SCN | 3.416027941 | 0.000635417 | ✓ |
| 31. Cingulum_Ant_L | DMN | 3.2382421 | 0.001202687 | ✓ |
| 33. Cingulum_Mid_L | SAL | 3.2382421 | 0.001202687 | ✓ |
| 32. Cingulum_Ant_R | DMN | 3.06045626 | 0.00221 | ✓ |
| 22. Olfactory_R | LIM | 3.035058282 | 0.002404892 | ✓ |
| 58. Postcentral_R | SOM | -3.009660305 | 0.0026154 | ✓ |
| 57. Postcentral_L | SOM | -2.984262328 | 0.002842629 | ✓ |
| 85. Temporal_Mid_L | DMN | 2.908068396 | 0.003636688 | ✓ |
| 30. Insula_R | SAL | 2.908068396 | 0.003636688 | ✓ |
| 34. Cingulum_Mid_R | SAL | 2.831874464 | 0.004627601 | ✓ |
| 28. Rectus_R | LIM | 2.831874464 | 0.004627601 | ✓ |
| 26. Frontal_Med_Orb_R | DMN | 2.806476487 | 0.005008657 | ✓ |
| 48. Lingual_R | VIS | -2.78107851 | 0.005417863 | ✓ |
| 44. Calcarine_R | VIS | -2.78107851 | 0.005417863 | ✓ |
| 46. Cuneus_R | VIS | -2.679486601 | 0.007373515 | ✓ |
| 2. Precentral_R | SOM | -2.679486601 | 0.007373515 | ✓ |
| 43. Calcarine_L | VIS | -2.654088624 | 0.007952294 | ✓ |
| 49. Occipital_Sup_L | VIS | -2.577894692 | 0.009940429 | ✓ |
| 25. Frontal_Med_Orb_L | DMN | 2.527098737 | 0.011500916 | ✓ |
| 45. Cuneus_L | VIS | -2.527098737 | 0.011500916 | ✓ |
| 16. Frontal_Inf_Orb_R | DMN | 2.476302783 | 0.013275094 | ✓ |
| 56. Fusiform_R | VIS | -2.450904805 | 0.014249763 | ✓ |
| 81. Temporal_Sup_L | SOM | 2.400108851 | 0.016390197 | ✓ |
| 69. Paracentral_Lobule_L | SOM | -2.374710874 | 0.017562701 | x |
| 29, Insula_L | SAL | 2.349312896 | 0.018808094 | x |
| 83. Temporal_Pole_Sup_L | LIM | 2.349312896 | 0.018808094 | x |
| 79. Heschl_L | SOM | 2.323914919 | 0.020130054 | x |
| 82. Temporal_Sup_R | SOM | 2.323914919 | 0.020130054 | x |
| 50. Occipital_Sup_R | VIS | -2.323914919 | 0.020130054 | x |
| 66. Angular_R | DMN | 2.298516942 | 0.021532385 | x |
| 80. Heschl_R | SOM | 2.273118965 | 0.023019012 | x |
| 54. Occipital_Inf_R | VIS | -2.22232301 | 0.026261484 | x |
| 23. Frontal_Sup_Medial_L | DMN | 2.196925033 | 0.028025801 | x |
| 65. Angular_L | DMN | 2.146129078 | 0.031862682 | x |
| 42. Amygdala_R | SCN | 2.120731101 | 0.033944438 | x |
| 47. Lingual_L | VIS | -2.095333124 | 0.03614139 | x |

| | | | | |
|---|---|---|---|---|
| 70. Paracentral_Lobule_R | SOM | -2.095333124 | 0.03614139 | x |
| 52. Occipital_Mid_R | VIS | -2.031471838 | 0.042207154 | x |
| 27. Rectus_L | LIM | 1.993741215 | 0.046180343 | x |

***Supplementary Table S8. Trophic level change in MBCT+TAU rumination.*** *Brain areas significant change between after vs. before, with the assigned resting-state network in which it shows the greatest contribution, zvalue, p-value and if they survive correction by multiple comparisons (✓) or not (x).*

| **Brain area** | **RSN** | **Z-value** | **P-value** | **Correction** |
|---|---|---|---|---|
| 75. Pallidum_L | SCN | 2.380899023 | 0.017270445 | x |
| 46. Cuneus_R | VIS | -2.346141373 | 0.018968911 | x |
| 50. Occipital_Sup_R | VIS | -2.172353123 | 0.029829038 | x |
| 73. Putamen_L | SCN | 2.102837823 | 0.035479948 | x |
| 45. Cuneus_L | VIS | -2.033322523 | 0.042019949 | x |
| 47. Lingual_L | VIS | -1.963807223 | 0.049552452 | x |

***Supplementary Table S9. Trophic level change in TAU rumination.*** *Brain areas significant change between after vs. before, with the assigned resting-state network in which it shows the greatest contribution, z-value, p-value and if they survive correction by multiple comparisons (✓) or not (x).*

| | RUM MBCT+TAU BEFORE - asymptomatic | | | RUM MBCT+TAU AFTER - asymptomatic | | |
|---|---|---|---|---|---|---|
| Top 20% | 54. Occipital_Inf_R | VIS | 0.485 | 76. Pallidum_R | SCN | 0.429 |
| | 36. Cingulum_Post_R | DMN | 0.432 | 71. Caudate_L | SCN | 0.426 |
| | 49. Occipital_Sup_L | VIS | 0.420 | 72. Caudate_R | SCN | 0.409 |
| | 43. Calcarine_L | VIS | 0.420 | 78. Thalamus_R | SCN | 0.397 |
| | 48. Lingual_R | VIS | 0.419 | 77. Thalamus_L | SCN | 0.394 |
| | 67. Precuneus_L | DMN | 0.415 | 75. Pallidum_L | SCN | 0.386 |
| | 53. Occipital_Inf_L | VIS | 0.412 | 22. Olfactory_R | LIM | 0.372 |
| | 59. Parietal_Sup_L | DAN | 0.412 | 74. Putamen_R | SCN | 0.356 |
| | 35. Cingulum_Post_L | DMN | 0.412 | 36. Cingulum_Post_R | DMN | 0.334 |
| | 90. Temporal_Inf_R | LIM | 0.411 | 81. Temporal_Sup_L | SOM | 0.329 |
| | 50. Occipital_Sup_R | VIS | 0.408 | 83. Temporal_Pole_Sup_L | LIM | 0.324 |
| | 71. Caudate_L | SCN | 0.407 | 7. Frontal_Mid_L | CNT | 0.324 |
| | 45. Cuneus_L | VIS | 0.401 | 30. Insula_R | SAL | 0.324 |
| | 47. Lingual_L | VIS | 0.400 | 15. Frontal_Inf_Orb_L | DMN | 0.322 |
| | 9. Frontal_Mid_Orb_L | CNT | 0.397 | 73. Putamen_L | SCN | 0.321 |
| | 38. Hippocampus_R | SCN | 0.397 | 90. Temporal_Inf_R | LIM | 0.316 |
| | 44. Calcarine_R | VIS | 0.396 | 89. Temporal_Inf_L | LIM | 0.315 |
| | 60. Parietal_Sup_R | DAN | 0.395 | 53. Occipital_Inf_L | VIS | 0.309 |
| Bottom 20% | 79. Heschl_L | SOM | 0.257 | 24. Frontal_Sup_Medial_R | DMN | 0.183 |
| | 32. Cingulum_Ant_R | DMN | 0.252 | 70. Paracentral_Lobule_R | SOM | 0.177 |
| | 66. Angular_R | DMN | 0.241 | 64. SupraMarginal_R | SAL | 0.175 |
| | 30. Insula_R | SAL | 0.237 | 49. Occipital_Sup_L | VIS | 0.172 |
| | 42. Amygdala_R | SCN | 0.236 | 1. Precentral_L | SOM | 0.169 |
| | 22. Olfactory_R | LIM | 0.232 | 86. Temporal_Mid_R | DMN | 0.166 |
| | 12. Frontal_Inf_Oper_R | CNT | 0.231 | 52. Occipital_Mid_R | VIS | 0.164 |
| | 63. SupraMarginal_L | SAL | 0.230 | 63. SupraMarginal_L | SAL | 0.163 |
| | 78. Thalamus_R | SCN | 0.227 | 88. Temporal_Pole_Mid_R | LIM | 0.163 |
| | 14. Frontal_Inf_Tri_R | CNT | 0.221 | 50. Occipital_Sup_R | VIS | 0.160 |
| | 77. Thalamus_L | SCN | 0.217 | 41. Amygdala_L | SCN | 0.159 |
| | 73. Putamen_L | SCN | 0.207 | 69. Paracentral_Lobule_L | SOM | 0.151 |
| | 64. SupraMarginal_R | SAL | 0.198 | 45. Cuneus_L | VIS | 0.149 |
| | 74. Putamen_R | SCN | 0.188 | 4. Frontal_Sup_R | DMN | 0.146 |
| | 75. Pallidum_L | SCN | 0.183 | 46. Cuneus_R | VIS | 0.127 |
| | 29, Insula_L | SAL | 0.165 | 57. Postcentral_L | SOM | 0.111 |
| | 76. Pallidum_R | SCN | 0.151 | 2. Precentral_R | SOM | 0.105 |
| | 41. Amygdala_L | SCN | 0.135 | 58. Postcentral_R | SOM | 0.088 |

***Supplementary Table S10. Trophic levels of each session for MBCT+TAU treatment during rumination in the trajectory from asymptomatic to asymptomatic.*** *The table shows the brain areas with highest 20% (first row) and lowest 20% trophic levels before (left column) and after (right column) treatment. Each brain area is assigned the resting-state network in which it shows the greatest contribution, and its trophic level.*

| | RUM MBCT+TAU BEFORE - symptomatic | | | RUM MBCT+TAU AFTER - asymptomatic | | |
|---|---|---|---|---|---|---|
| Top 20% | 60. Parietal_Sup_R | DAN | 0.416 | 72. Caudate_R | SCN | 0.759 |
| | 36. Cingulum_Post_R | DMN | 0.409 | 73. Putamen_L | SCN | 0.703 |
| | 67. Precuneus_L | DMN | 0.404 | 71. Caudate_L | SCN | 0.697 |
| | 59. Parietal_Sup_L | DAN | 0.395 | 74. Putamen_R | SCN | 0.684 |
| | 68. Precuneus_R | DMN | 0.384 | 76. Pallidum_R | SCN | 0.667 |
| | 54. Occipital_Inf_R | VIS | 0.383 | 78. Thalamus_R | SCN | 0.664 |
| | 50. Occipital_Sup_R | VIS | 0.370 | 77. Thalamus_L | SCN | 0.652 |
| | 43. Calcarine_L | VIS | 0.367 | 75. Pallidum_L | SCN | 0.642 |
| | 53. Occipital_Inf_L | VIS | 0.362 | 35. Cingulum_Post_L | DMN | 0.573 |
| | 9. Frontal_Mid_Orb_L | CNT | 0.360 | 36. Cingulum_Post_R | DMN | 0.570 |
| | 72. Caudate_R | SCN | 0.346 | 65. Angular_L | DMN | 0.503 |
| | 5. Frontal_Sup_Orb_L | LIM | 0.346 | 66. Angular_R | DMN | 0.497 |
| | 51. Occipital_Mid_L | VIS | 0.345 | 31. Cingulum_Ant_L | DMN | 0.485 |
| | 48. Lingual_R | VIS | 0.345 | 32. Cingulum_Ant_R | DMN | 0.484 |
| | 84. Temporal_Pole_Sup_R | LIM | 0.343 | 67. Precuneus_L | DMN | 0.477 |
| | 7. Frontal_Mid_L | CNT | 0.342 | 23. Frontal_Sup_Medial_L | DMN | 0.471 |
| | 52. Occipital_Mid_R | VIS | 0.338 | 24. Frontal_Sup_Medial_R | DMN | 0.459 |
| | 82. Temporal_Sup_R | SOM | 0.337 | 7. Frontal_Mid_L | CNT | 0.450 |
| Bottom 20% | 25. Frontal_Med_Orb_L | DMN | 0.248 | 43. Calcarine_L | VIS | 0.259 |
| | 28. Rectus_R | LIM | 0.241 | 50. Occipital_Sup_R | VIS | 0.251 |
| | 15. Frontal_Inf_Orb_L | DMN | 0.241 | 56. Fusiform_R | VIS | 0.250 |
| | 31. Cingulum_Ant_L | DMN | 0.239 | 48. Lingual_R | VIS | 0.237 |
| | 40. ParaHippocampal_R | LIM | 0.236 | 17. Rolandic_Oper_L | SOM | 0.235 |
| | 22. Olfactory_R | LIM | 0.236 | 18. Rolandic_Oper_R | SOM | 0.230 |
| | 16. Frontal_Inf_Orb_R | DMN | 0.231 | 70. Paracentral_Lobule_R | SOM | 0.229 |
| | 83. Temporal_Pole_Sup_L | LIM | 0.216 | 49. Occipital_Sup_L | VIS | 0.229 |
| | 32. Cingulum_Ant_R | DMN | 0.213 | 47. Lingual_L | VIS | 0.226 |
| | 30. Insula_R | SAL | 0.208 | 20. Supp_Motor_Area_R | SOM | 0.221 |
| | 74. Putamen_R | SCN | 0.204 | 44. Calcarine_R | VIS | 0.213 |
| | 39. ParaHippocampal_L | LIM | 0.202 | 45. Cuneus_L | VIS | 0.204 |
| | 73. Putamen_L | SCN | 0.195 | 46. Cuneus_R | VIS | 0.181 |
| | 88. Temporal_Pole_Mid_R | LIM | 0.186 | 69. Paracentral_Lobule_L | SOM | 0.176 |
| | 41. Amygdala_L | SCN | 0.183 | 1. Precentral_L | SOM | 0.157 |
| | 42. Amygdala_R | SCN | 0.182 | 58. Postcentral_R | SOM | 0.152 |
| | 87. Temporal_Pole_Mid_L | LIM | 0.181 | 57. Postcentral_L | SOM | 0.141 |
| | 29, Insula_L | SAL | 0.165 | 2. Precentral_R | SOM | 0.131 |

***Supplementary Table S11. Trophic levels of each session for MBCT+TAU treatment during rumination in the trajectory from symptomatic to asymptomatic.*** *The table shows the brain areas with highest 20% (first row) and lowest 20% trophic levels before (left column) and after (right column) treatment. Each brain area is assigned the resting-state network in which it shows the greatest contribution, and its trophic level.*

|  | RUM MBCT+TAU BEFORE - symptomatic |  |  | RUM MBCT+TAU AFTER - symptomatic |  |  |
|---|---|---|---|---|---|---|
| Top 20% | 43. Calcarine_L | VIS | 0.399 | 72. Caudate_R | SCN | 0.564 |
| | 67. Precuneus_L | DMN | 0.398 | 78. Thalamus_R | SCN | 0.513 |
| | 49. Occipital_Sup_L | VIS | 0.390 | 82. Temporal_Sup_R | SOM | 0.504 |
| | 44. Calcarine_R | VIS | 0.387 | 81. Temporal_Sup_L | SOM | 0.499 |
| | 68. Precuneus_R | DMN | 0.384 | 77. Thalamus_L | SCN | 0.498 |
| | 45. Cuneus_L | VIS | 0.376 | 80. Heschl_R | SOM | 0.497 |
| | 70. Paracentral_Lobule_R | SOM | 0.370 | 76. Pallidum_R | SCN | 0.488 |
| | 53. Occipital_Inf_L | VIS | 0.369 | 71. Caudate_L | SCN | 0.485 |
| | 54. Occipital_Inf_R | VIS | 0.369 | 79. Heschl_L | SOM | 0.462 |
| | 36. Cingulum_Post_R | DMN | 0.368 | 75. Pallidum_L | SCN | 0.449 |
| | 48. Lingual_R | VIS | 0.364 | 22. Olfactory_R | LIM | 0.445 |
| | 59. Parietal_Sup_L | DAN | 0.358 | 67. Precuneus_L | DMN | 0.442 |
| | 46. Cuneus_R | VIS | 0.356 | 28. Rectus_R | LIM | 0.440 |
| | 72. Caudate_R | SCN | 0.354 | 68. Precuneus_R | DMN | 0.439 |
| | 51. Occipital_Mid_L | VIS | 0.352 | 36. Cingulum_Post_R | DMN | 0.427 |
| | 50. Occipital_Sup_R | VIS | 0.352 | 27. Rectus_L | LIM | 0.425 |
| | 60. Parietal_Sup_R | DAN | 0.352 | 85. Temporal_Mid_L | DMN | 0.424 |
| | 35. Cingulum_Post_L | DMN | 0.351 | 33. Cingulum_Mid_L | SAL | 0.389 |
| Bottom 20% | 27. Rectus_L | LIM | 0.248 | 58. Postcentral_R | SOM | 0.301 |
| | 64. SupraMarginal_R | SAL | 0.246 | 37. Hippocampus_L | SCN | 0.293 |
| | 13. Frontal_Inf_Tri_L | CNT | 0.244 | 88. Temporal_Pole_Mid_R | LIM | 0.288 |
| | 1. Precentral_L | SOM | 0.243 | 39. ParaHippocampal_L | LIM | 0.280 |
| | 87. Temporal_Pole_Mid_L | LIM | 0.242 | 13. Frontal_Inf_Tri_L | CNT | 0.276 |
| | 14. Frontal_Inf_Tri_R | CNT | 0.241 | 15. Frontal_Inf_Orb_L | DMN | 0.275 |
| | 16. Frontal_Inf_Orb_R | DMN | 0.239 | 16. Frontal_Inf_Orb_R | DMN | 0.272 |
| | 25. Frontal_Med_Orb_L | DMN | 0.231 | 1. Precentral_L | SOM | 0.269 |
| | 88. Temporal_Pole_Mid_R | LIM | 0.228 | 2. Precentral_R | SOM | 0.268 |
| | 73. Putamen_L | SCN | 0.227 | 21. Olfactory_L | LIM | 0.267 |
| | 28. Rectus_R | LIM | 0.227 | 42. Amygdala_R | SCN | 0.265 |
| | 11. Frontal_Inf_Oper_L | CNT | 0.226 | 14. Frontal_Inf_Tri_R | CNT | 0.262 |
| | 31. Cingulum_Ant_L | DMN | 0.223 | 57. Postcentral_L | SOM | 0.247 |
| | 30. Insula_R | SAL | 0.216 | 40. ParaHippocampal_R | LIM | 0.240 |
| | 42. Amygdala_R | SCN | 0.211 | 11. Frontal_Inf_Oper_L | CNT | 0.234 |
| | 26. Frontal_Med_Orb_R | DMN | 0.210 | 29, Insula_L | SAL | 0.231 |
| | 74. Putamen_R | SCN | 0.209 | 41. Amygdala_L | SCN | 0.205 |
| | 29, Insula_L | SAL | 0.206 | 87. Temporal_Pole_Mid_L | LIM | 0.199 |

***Supplementary Table S12. Trophic levels of each session for MBCT+TAU treatment during rumination in the trajectory from symptomatic to symptomatic.*** *The table shows the brain areas with highest 20% (first row) and lowest 20% trophic levels before (left column) and after (right column) treatment. Each brain area is assigned the resting-state network in which it shows the greatest contribution, and its trophic level.*

| | MBCT+TAU | | | | | TAU | | | | | MBCT+TAU & TAU | | | | |
|---|---|---|---|---|---|---|---|---|---|---|---|---|---|---|---|
| | Spearman | | Partial Spearman | | N | Spearman | | Partial Spearman | | N | Spearman | | Partial Spearman | | N |
| | Rho | P-val | Rho | P-val | | Rho | P-val | Rho | P-val | | Rho | P-val | Rho | P-val | |
| **Mental health outcomes** | | | | | | | | | | | | | | | |
| **QIDS POST** | -0.312 | 0.121 | **-0.481** | **0.023** | 26 | -0.068 | 0.783 | 0.055 | 0.846 | 19 | -0.273 | 0.070 | **-0.320** | **0.041** | 45 |
| **QIDS 3M** | -0.315 | 0.118 | -0.398 | 0.067 | 26 | -0.283 | 0.270 | -0.244 | 0.423 | 17 | **-0.345** | **0.024** | **-0.344** | **0.032** | 43 |
| **PSS** | **-0.505** | **0.010** | **-0.562** | **0.008** | 25 | -0.396 | 0.103 | -0.288 | 0.341 | 17 | **-0.457** | **0.002** | **-0.494** | **0.002** | 42 |
| **Mechanism outcomes** | | | | | | | | | | | | | | | |
| **FFMQ** | **0.448** | **0.025** | **0.529** | **0.014** | 25 | 0.153 | 0.545 | 0.117 | 0.705 | 17 | **0.382** | **0.011** | **0.420** | **0.009** | 42 |
| **RRS** | **-0.471** | **0.017** | **-0.509** | **0.018** | 25 | -0.058 | 0.820 | -0.049 | 0.874 | 17 | **-0.305** | **0.047** | **-0.355** | **0.029** | 42 |
| **EQ** | **0.645** | **<0.001** | **0.761** | **<0.001** | 25 | 0.032 | 0.899 | 0.083 | 0.787 | 17 | **0.485** | **0.001** | **0.536** | **0.001** | 42 |
| **MAIA_NO** | 0.210 | 0.313 | 0.281 | 0.217 | 25 | -0.331 | 0.179 | -0.450 | 0.123 | 17 | 0.164 | 0.295 | 0.183 | 0.273 | 42 |
| **MAIA_ND** | -0.266 | 0.199 | -0.308 | 0.174 | 25 | 0.049 | 0.847 | 0.033 | 0.914 | 17 | -0.213 | 0.170 | -0.271 | 0.100 | 42 |
| **MAIA_EA** | 0.303 | 0.141 | 0.408 | 0.066 | 25 | 0.240 | 0.338 | 0.412 | 0.162 | 17 | **0.372** | **0.014** | **0.426** | **0.008** | 42 |
| **MAIA_AR** | **0.609** | **0.001** | **0.660** | **0.001** | 25 | -0.163 | 0.519 | -0.307 | 0.307 | 17 | **0.430** | **0.004** | **0.431** | **0.007** | 42 |
| **MAIA_BL** | 0.360 | 0.077 | **0.471** | **0.031** | 25 | -0.090 | 0.721 | -0.036 | 0.908 | 17 | 0.275 | 0.074 | **0.329** | **0.044** | 42 |
| **MAIA_TR** | 0.214 | 0.303 | 0.228 | 0.321 | 25 | -0.067 | 0.791 | 0.057 | 0.852 | 17 | 0.135 | 0.387 | 0.152 | 0.364 | 42 |

***Supplementary Table S13. Correlation between change (post-pre) scores in clinical and behavioural outcomes and change (post-pre) in breadth of hierarchy for rumination.*** *Each row is a clinical and behavioural outcome with the following abbreviations: QIDS, Quick Inventory of Depressive Symptomatology, post-treatment (post) and 3 months after treatment (3M); Perceived Stress Scale (PSS); Five Factor Mindfulness Questionnaire (FFMQ); Ruminative Response Scale (RRS), Total; Experience Questionnaire ( EQ) for decentering; Multidimensional Assessment of Interoceptive Awareness (MAIA) for noticing (NO), not-distracting (ND), emotional awareness (EA), attention regulation (AR), body listening (BL) and trusting (TR). Each column corresponds to a treatment (MBCT+TAU, TAU, or MBCT+TAU and TAU altogether), as well as strength and significance for Spearman and Partial Spearman correlations corrected by age, sex, ADM and baseline QIDS, and number of individuals (N). In bold and green the significant correlations.*

| | MBCT+TAU | | | | | TAU | | | | | MBCT+TAU & TAU | | | | |
|---|---|---|---|---|---|---|---|---|---|---|---|---|---|---|---|
| | Spearman | | Partial Spearman | | N | Spearman | | Partial Spearman | | N | Spearman | | Partial Spearman | | N |
| | Rho | P-val | Rho | P-val | | Rho | P-val | Rho | P-val | | Rho | P-val | Rho | P-val | |
| **Mental health outcomes** | | | | | | | | | | | | | | | |
| **QIDS** | -0.081 | 0.694 | -0.098 | 0.656 | 26 | 0.231 | 0.341 | 0.380 | 0.146 | 19 | -0.031 | 0.840 | -0.039 | 0.804 | 45 |
| **PSS** | 0.032 | 0.881 | 0.073 | 0.752 | 25 | 0.296 | 0.193 | 0.347 | 0.205 | 19 | 0.102 | 0.502 | 0.108 | 0.509 | 44 |
| **Mechanism outcomes** | | | | | | | | | | | | | | | |
| **FFMQ** | -0.160 | 0.445 | -0.167 | 0.469 | 25 | -0.390 | 0.080 | **-0.525** | **0.045** | 19 | -0.272 | 0.067 | -0.293 | 0.066 | 44 |
| **RRS** | 0.120 | 0.569 | 0.167 | 0.470 | 25 | -0.014 | 0.951 | 0.081 | 0.775 | 19 | 0.037 | 0.807 | 0.054 | 0.740 | 44 |
| **EQ** | **-0.497** | **0.011** | **-0.588** | **0.005** | 25 | -0.378 | 0.091 | -0.509 | 0.053 | 19 | **-0.421** | **0.004** | **-0.504** | **0.001** | 44 |
| **MAIA_NO** | -0.277 | 0.180 | -0.225 | 0.327 | 25 | 0.166 | 0.471 | 0.204 | 0.465 | 19 | -0.156 | 0.302 | -0.119 | 0.463 | 44 |
| **MAIA_ND** | 0.023 | 0.912 | 0.034 | 0.885 | 25 | **-0.455** | **0.038** | -0.243 | 0.384 | 19 | -0.070 | 0.644 | -0.029 | 0.860 | 44 |
| **MAIA_EA** | -0.161 | 0.443 | -0.140 | 0.545 | 25 | -0.163 | 0.479 | -0.398 | 0.142 | 19 | -0.219 | 0.144 | -0.261 | 0.104 | 44 |
| **MAIA_AR** | **-0.493** | **0.012** | **-0.436** | **0.048** | 25 | -0.096 | 0.680 | -0.060 | 0.832 | 19 | **-0.337** | **0.022** | -0.309 | 0.052 | 44 |
| **MAIA_BL** | **-0.445** | **0.026** | -0.408 | 0.066 | 25 | 0.358 | 0.111 | 0.423 | 0.116 | 19 | -0.045 | 0.766 | -0.075 | 0.645 | 44 |
| **MAIA_TR** | 0.010 | 0.963 | 0.022 | 0.923 | 25 | -0.245 | 0.284 | -0.356 | 0.193 | 19 | -0.063 | 0.677 | -0.073 | 0.654 | 44 |

***Supplementary Table S14. Correlation between baseline scores in clinical and behavioural outcomes and change (post-pre) in breadth of hierarchy for rumination.*** *Each row is a clinical and behavioural outcomes with the following abbreviations: QIDS, Quick Inventory of Depressive Symptomatology, Perceived Stress Scale (PSS); Five Factor Mindfulness Questionnaire (FFMQ); Ruminative Response Scale (RRS); Experience Questionnaire ( EQ) for decentering; Multidimensional Assessment of Interoceptive Awareness (MAIA) for noticing (NO), not-distracting (ND), emotional awareness (EA), attention regulation (AR), body listening (BL) and trusting (TR). Each column corresponds to a treatment (MBCT+TAU, TAU, or MBCT+TAU and TAU altogether), as well as strength and significance for Spearman and Partial Spearman correlations corrected by age, sex, and ADM and baseline QIDS for all measures except for QIDS, and number of individuals (N). In bold and green the significant correlations.*

| | | Treatment effect on change in clinical or behavioural scores via change in brain marker | | | | | | | | | | | |
|---|---|---|---|---|---|---|---|---|---|---|---|---|---|
| | | QIDS POST | QIDS 3M | PSS | FFMQ | RRS | EQ | MAIA_NO | MAIA_ND | MAIA_EA | MAIA_AR | MAIA_BL | MAIA_TR |
| Non-adjusted | **p-vals** | | | | | | | | | | | | |
| | **p-val a** | 0.015 | 0.026 | 0.015 | 0.013 | 0.021 | 0.018 | 0.022 | 0.018 | 0.018 | 0.018 | 0.015 | 0.017 |
| | **p-val b** | 0.147 | 0.037 | 0.000 | 0.009 | 0.018 | 0.004 | 0.573 | 0.242 | 0.049 | 0.025 | 0.300 | 0.242 |
| | **p-val ab** | 0.183 | 0.087 | 0.011 | 0.024 | 0.039 | 0.025 | 0.457 | 0.413 | 0.028 | 0.034 | 0.221 | 0.184 |
| | **p-val c** | 0.009 | 0.027 | 0.027 | 0.031 | 0.754 | 0.000 | 0.001 | 0.191 | 0.000 | 0.000 | 0.000 | 0.128 |
| | **p-val cp** | 0.073 | 0.145 | 0.272 | 0.187 | 0.703 | 0.001 | 0.005 | 0.493 | 0.003 | 0.002 | 0.000 | 0.236 |
| | **strength** | | | | | | | | | | | | |
| | **t a** | 0.198 | 0.189 | 0.199 | 0.199 | 0.199 | 0.199 | 0.199 | 0.199 | 0.199 | 0.199 | 0.199 | 0.199 |
| | **t b** | -3.490 | -4.577 | -9.505 | 9.999 | -12.369 | 8.342 | 0.916 | -2.015 | 4.264 | 6.378 | 1.027 | 1.351 |
| | **t ab** | -0.691 | -0.866 | -1.890 | 1.988 | -2.459 | 1.659 | 0.182 | -0.401 | 0.848 | 1.268 | 0.204 | 0.269 |
| | **t c** | -3.798 | -3.097 | -4.320 | 5.273 | -0.938 | 8.636 | 3.171 | -1.042 | 4.582 | 6.084 | 2.700 | 1.187 |
| | **t cp** | -3.106 | -2.232 | -2.430 | 3.285 | 1.521 | 6.977 | 2.989 | -0.642 | 3.735 | 4.816 | 2.496 | 0.918 |
| | **lower CI** | | | | | | | | | | | | |
| | **CI_L a** | 0.040 | 0.024 | 0.037 | 0.038 | 0.033 | 0.033 | 0.035 | 0.035 | 0.034 | 0.033 | 0.040 | 0.029 |
| | **CI_L b** | -8.444 | -9.775 | -15.568 | 2.986 | -23.669 | 2.839 | -2.581 | -6.194 | 0.027 | 0.832 | -0.895 | -1.004 |
| | **CI_L ab** | -2.497 | -2.726 | -4.606 | 0.156 | -6.880 | 0.121 | -0.430 | -1.703 | 0.088 | 0.061 | -0.111 | -0.113 |
| | **CI_L c** | -6.751 | -5.852 | -8.030 | 0.532 | -7.382 | 4.872 | 1.274 | -2.629 | 2.004 | 3.494 | 1.557 | -0.345 |
| | **CI_L cp** | -6.172 | -5.004 | -6.162 | -1.800 | -5.588 | 3.099 | 0.923 | -2.433 | 1.360 | 1.813 | 1.295 | -0.662 |
| | **upper CI** | | | | | | | | | | | | |
| | **CI_H a** | 0.347 | 0.345 | 0.352 | 0.353 | 0.353 | 0.349 | 0.355 | 0.352 | 0.353 | 0.351 | 0.350 | 0.348 |
| | **CI_H b** | 1.589 | -0.001 | -3.442 | 17.878 | -2.012 | 14.737 | 4.779 | 1.362 | 8.568 | 11.846 | 3.124 | 3.991 |
| | **CI_H ab** | 0.169 | -0.003 | -0.195 | 4.938 | -0.130 | 4.029 | 1.339 | 0.175 | 2.332 | 3.398 | 0.889 | 1.170 |
| | **CI_H c** | -0.871 | -0.436 | -0.438 | 9.333 | 6.903 | 12.236 | 5.105 | 0.600 | 7.353 | 8.925 | 3.875 | 2.832 |
| | **CI_H cp** | -0.008 | 0.514 | 1.171 | 7.576 | 9.644 | 10.670 | 5.131 | 1.424 | 6.301 | 8.112 | 3.675 | 2.461 |
| Adjusted | **p-vals** | | | | | | | | | | | | |
| | **p-val a** | 0.020 | 0.065 | 0.022 | 0.033 | 0.023 | 0.028 | 0.026 | 0.024 | 0.021 | 0.024 | 0.025 | 0.018 |
| | **p-val b** | 0.085 | 0.088 | 0.000 | 0.042 | 0.031 | 0.133 | 0.645 | 0.018 | 0.059 | 0.216 | 0.238 | 0.396 |
| | **p-val ab** | 0.092 | 0.142 | 0.009 | 0.056 | 0.041 | 0.123 | 0.523 | 0.059 | 0.045 | 0.171 | 0.178 | 0.314 |
| | **p-val c** | 0.008 | 0.010 | 0.009 | 0.114 | 0.334 | 0.001 | 0.009 | 0.504 | 0.001 | 0.001 | 0.000 | 0.061 |
| | **p-val cp** | 0.083 | 0.064 | 0.262 | 0.350 | 0.793 | 0.002 | 0.021 | 0.508 | 0.012 | 0.002 | 0.001 | 0.132 |
| | **strength** | | | | | | | | | | | | |
| | **t a** | 0.194 | 0.171 | 0.190 | 0.178 | 0.194 | 0.169 | 0.187 | 0.196 | 0.187 | 0.184 | 0.191 | 0.194 |
| | **t b** | -4.169 | -4.498 | -10.849 | 7.741 | -12.856 | 5.100 | 0.714 | -2.972 | 4.419 | 2.892 | 1.277 | 1.001 |
| | **t ab** | -0.809 | -0.770 | -2.061 | 1.377 | -2.499 | 0.861 | 0.134 | -0.583 | 0.828 | 0.531 | 0.244 | 0.195 |
| | **t c** | -4.042 | -3.278 | -4.171 | 4.173 | -1.857 | 7.529 | 2.544 | -0.208 | 4.370 | 5.542 | 2.740 | 1.446 |
| | **t cp** | -3.233 | -2.508 | -2.110 | 2.797 | 0.642 | 6.668 | 2.410 | 0.376 | 3.542 | 5.011 | 2.496 | 1.251 |
| | **lower CI** | | | | | | | | | | | | |
| | **CI_L a** | 0.134 | 0.105 | 0.135 | 0.125 | 0.138 | 0.116 | 0.132 | 0.138 | 0.134 | 0.128 | 0.134 | 0.141 |
| | **CI_L b** | -5.843 | -6.676 | -13.147 | 5.373 | -17.177 | 3.032 | -0.287 | -3.767 | 2.818 | 1.374 | 0.526 | 0.207 |
| | **CI_L ab** | -1.355 | -1.381 | -3.055 | 0.785 | -4.063 | 0.429 | -0.004 | -0.882 | 0.510 | 0.250 | 0.105 | 0.059 |
| | **CI_L c** | -5.098 | -4.082 | -5.185 | 2.092 | -3.746 | 6.100 | 1.885 | -0.587 | 3.308 | 4.608 | 2.243 | 0.903 |
| | **CI_L cp** | -4.336 | -3.372 | -3.450 | 0.699 | -1.489 | 5.144 | 1.726 | -0.011 | 2.498 | 3.939 | 2.041 | 0.667 |
| | **upper CI** | | | | | | | | | | | | |
| | **CI_H a** | 0.240 | 0.217 | 0.243 | 0.230 | 0.245 | 0.219 | 0.242 | 0.249 | 0.237 | 0.237 | 0.243 | 0.246 |
| | **CI_H b** | -2.745 | -3.101 | -8.317 | 10.322 | -9.013 | 7.366 | 1.566 | -2.094 | 6.081 | 4.601 | 1.881 | 1.832 |
| | **CI_H ab** | -0.440 | -0.466 | -1.299 | 2.269 | -1.481 | 1.561 | 0.360 | -0.384 | 1.353 | 1.070 | 0.452 | 0.438 |
| | **CI_H c** | -3.163 | -2.438 | -2.635 | 5.127 | 1.119 | 8.371 | 2.959 | 0.340 | 5.140 | 6.257 | 3.078 | 1.809 |
| | **CI_H cp** | -2.267 | -1.592 | -0.757 | 3.942 | 3.756 | 7.734 | 2.915 | 0.905 | 4.327 | 5.835 | 2.880 | 1.699 |

***Supplementary Table S15. Mediation analysis of treatment (TAU vs. MBCT+TAU) on change clinical and behavioural outcomes via change in breadth of hierarchy for rumination, and adjusted by age, sex, ADM and baseline QIDS.*** *Significant mediations are coloured in green. Each column is a clinical and behavioural outcomes with the following abbreviations: QIDS, Quick Inventory of Depressive Symptomatology, post-treatment (post) and 3 months after treatment (3M); Perceived Stress Scale (PSS); Five Factor Mindfulness Questionnaire (FFMQ); Ruminative Response Scale (RRS); Experience Questionnaire ( EQ) for decentering; Multidimensional Assessment of Interoceptive Awareness (MAIA) for noticing (NO), not-distracting (ND), emotional awareness (EA), attention regulation (AR), body listening (BL) and trusting (TR). In green the significant results.*

| | | Treatment effect on change in brain marker via change in clinical or behavioural scores | | | | | | | | | | |
|---|---|---|---|---|---|---|---|---|---|---|---|---|
| | | QIDS POST | PSS | FFMQ | RRS | EQ | MAIA_NO | MAIA_ND | MAIA_EA | MAIA_AR | MAIA_BL | MAIA_TR |
| Non-adjusted | **p-vals** | | | | | | | | | | | |
| | **p-val a** | 0.012 | 0.025 | 0.029 | 0.792 | 0.000 | 0.002 | 0.204 | 0.000 | 0.000 | 0.000 | 0.127 |
| | **p-val b** | 0.150 | 0.001 | 0.005 | 0.011 | 0.003 | 0.600 | 0.263 | 0.039 | 0.011 | 0.284 | 0.250 |
| | **p-val ab** | 0.115 | 0.027 | 0.021 | 0.706 | 0.005 | 0.513 | 0.181 | 0.031 | 0.006 | 0.243 | 0.194 |
| | **p-val c** | 0.015 | 0.015 | 0.015 | 0.016 | 0.020 | 0.013 | 0.015 | 0.018 | 0.020 | 0.015 | 0.019 |
| | **p-val cp** | 0.040 | 0.123 | 0.094 | 0.020 | 0.400 | 0.039 | 0.033 | 0.189 | 0.323 | 0.094 | 0.028 |
| | **strength** | | | | | | | | | | | |
| | **t a** | -3.798 | -4.320 | 5.273 | -0.938 | 8.636 | 3.171 | -1.042 | 4.582 | 6.084 | 2.700 | 1.187 |
| | **t b** | -0.010 | -0.017 | 0.013 | -0.007 | 0.015 | 0.005 | -0.017 | 0.016 | 0.019 | 0.018 | 0.012 |
| | **t ab** | 0.037 | 0.073 | 0.067 | 0.006 | 0.129 | 0.017 | 0.018 | 0.073 | 0.114 | 0.050 | 0.014 |
| | **t c** | 0.198 | 0.199 | 0.199 | 0.199 | 0.199 | 0.199 | 0.199 | 0.199 | 0.199 | 0.199 | 0.199 |
| | **t cp** | 0.161 | 0.126 | 0.132 | 0.193 | 0.070 | 0.182 | 0.181 | 0.126 | 0.084 | 0.149 | 0.185 |
| | **lower CI** | | | | | | | | | | | |
| | **CI_L a** | -6.756 | -8.169 | 0.633 | -7.367 | 4.949 | 1.238 | -2.600 | 2.054 | 3.334 | 1.526 | -0.360 |
| | **CI_L b** | -0.025 | -0.028 | 0.005 | -0.012 | 0.006 | -0.015 | -0.040 | 0.001 | 0.005 | -0.020 | -0.011 |
| | **CI_L ab** | -0.007 | 0.006 | 0.008 | -0.045 | 0.039 | -0.043 | -0.011 | 0.006 | 0.034 | -0.043 | -0.005 |
| | **CI_L c** | 0.038 | 0.039 | 0.043 | 0.035 | 0.037 | 0.038 | 0.037 | 0.035 | 0.033 | 0.042 | 0.034 |
| | **CI_L cp** | 0.007 | -0.036 | -0.021 | 0.029 | -0.104 | 0.008 | 0.014 | -0.059 | -0.090 | -0.023 | 0.023 |
| | **upper CI** | | | | | | | | | | | |
| | **CI_H a** | -0.873 | -0.442 | 9.357 | 6.724 | 12.285 | 5.105 | 0.634 | 7.445 | 8.850 | 3.862 | 2.821 |
| | **CI_H b** | 0.004 | -0.007 | 0.022 | -0.001 | 0.026 | 0.025 | 0.016 | 0.032 | 0.033 | 0.050 | 0.031 |
| | **CI_H ab** | 0.141 | 0.185 | 0.171 | 0.067 | 0.254 | 0.094 | 0.086 | 0.176 | 0.244 | 0.163 | 0.076 |
| | **CI_H c** | 0.342 | 0.355 | 0.355 | 0.352 | 0.352 | 0.356 | 0.350 | 0.353 | 0.351 | 0.356 | 0.350 |
| | **CI_H cp** | 0.300 | 0.274 | 0.270 | 0.341 | 0.227 | 0.341 | 0.342 | 0.305 | 0.236 | 0.319 | 0.337 |
| Adjusted | **p-vals** | | | | | | | | | | | |
| | **p-val a** | 0.010 | 0.008 | 0.110 | 0.339 | 0.001 | 0.012 | 0.542 | 0.001 | 0.001 | 0.000 | 0.069 |
| | **p-val b** | 0.085 | 0.001 | 0.056 | 0.023 | 0.162 | 0.695 | 0.008 | 0.069 | 0.221 | 0.263 | 0.414 |
| | **p-val ab** | 0.061 | 0.024 | 0.100 | 0.568 | 0.131 | 0.625 | 0.776 | 0.057 | 0.186 | 0.256 | 0.311 |
| | **p-val c** | 0.025 | 0.021 | 0.030 | 0.023 | 0.035 | 0.027 | 0.016 | 0.022 | 0.023 | 0.028 | 0.022 |
| | **p-val cp** | 0.094 | 0.152 | 0.105 | 0.029 | 0.300 | 0.045 | 0.011 | 0.209 | 0.178 | 0.186 | 0.045 |
| | **strength** | | | | | | | | | | | |
| | **t a** | -4.042 | -4.171 | 4.173 | -1.857 | 7.529 | 2.544 | -0.208 | 4.370 | 5.542 | 2.740 | 1.446 |
| | **t b** | -0.013 | -0.020 | 0.011 | -0.007 | 0.012 | 0.008 | -0.046 | 0.018 | 0.013 | 0.027 | 0.015 |
| | **t ab** | 0.052 | 0.085 | 0.047 | 0.012 | 0.087 | 0.020 | 0.009 | 0.079 | 0.073 | 0.073 | 0.022 |
| | **t c** | 0.194 | 0.190 | 0.178 | 0.194 | 0.169 | 0.187 | 0.196 | 0.187 | 0.184 | 0.191 | 0.194 |
| | **t cp** | 0.142 | 0.105 | 0.131 | 0.182 | 0.082 | 0.167 | 0.187 | 0.108 | 0.110 | 0.118 | 0.172 |
| | **lower CI** | | | | | | | | | | | |
| | **CI_L a** | -5.081 | -5.256 | 2.244 | -3.715 | 6.084 | 1.888 | -0.595 | 3.330 | 4.618 | 2.279 | 0.876 |
| | **CI_L b** | -0.018 | -0.025 | 0.007 | -0.009 | 0.006 | -0.005 | -0.056 | 0.011 | 0.006 | 0.011 | 0.003 |
| | **CI_L ab** | 0.031 | 0.055 | 0.023 | -0.002 | 0.047 | -0.008 | -0.011 | 0.049 | 0.039 | 0.027 | 0.008 |
| | **CI_L c** | 0.133 | 0.134 | 0.121 | 0.135 | 0.116 | 0.131 | 0.145 | 0.133 | 0.132 | 0.132 | 0.142 |
| | **CI_L cp** | 0.085 | 0.059 | 0.082 | 0.129 | 0.032 | 0.119 | 0.143 | 0.051 | 0.057 | 0.061 | 0.120 |
| | **upper CI** | | | | | | | | | | | |
| | **CI_H a** | -3.123 | -2.692 | 5.184 | 1.014 | 8.377 | 2.938 | 0.335 | 5.087 | 6.264 | 3.107 | 1.782 |
| | **CI_H b** | -0.008 | -0.016 | 0.014 | -0.005 | 0.016 | 0.019 | -0.032 | 0.023 | 0.021 | 0.038 | 0.029 |
| | **CI_H ab** | 0.087 | 0.119 | 0.074 | 0.029 | 0.132 | 0.051 | 0.029 | 0.112 | 0.121 | 0.110 | 0.052 |
| | **CI_H c** | 0.243 | 0.241 | 0.227 | 0.241 | 0.219 | 0.243 | 0.254 | 0.241 | 0.238 | 0.242 | 0.248 |
| | **CI_H cp** | 0.189 | 0.158 | 0.184 | 0.232 | 0.136 | 0.234 | 0.245 | 0.171 | 0.164 | 0.184 | 0.228 |

***Supplementary Table S16. Mediation analysis of treatment (TAU vs. MBCT+TAU) on change in breadth of hierarchy via change in clinical and behavioural outcomes for rumination, and adjusted by age, sex, ADM and baseline QIDS.*** *Significant mediations are coloured in green. Each column is a clinical and behavioural outcomes with the following abbreviations: QIDS, Quick Inventory of Depressive Symptomatology, post-treatment (post) and 3 months after treatment (3M); Perceived Stress Scale (PSS); Five Factor Mindfulness Questionnaire (FFMQ); Ruminative Response Scale (RRS); Experience Questionnaire ( EQ) for decentering; Multidimensional Assessment of Interoceptive Awareness (MAIA) for noticing (NO), not-distracting (ND), emotional awareness (EA), attention regulation (AR), body listening (BL) and trusting (TR). In green the significant results.*
